# Rare coding variants in *CHRNB2* reduce the likelihood of smoking

**DOI:** 10.1101/2022.10.27.22281470

**Authors:** Veera M. Rajagopal, Kyoko Watanabe, Joelle Mbatchou, Ariane Ayer, Peter Quon, Deepika Sharma, Michael D. Kessler, Kavita Praveen, Sahar Gelfman, Neelroop Parikshak, Jacqueline M. Otto, Suyin Bao, Shek Man Chim, Elias Pavlopoulos, Andreja Avbersek, Manav Kapoor, Esteban Chen, Marcus B. Jones, Michelle Leblanc, Jonathan Emberson, Rory Collins, Jason Torres, Pablo Kuri Morales, Roberto Tapia-Conyer, Jesus Alegre, Jaime Berumen, GHS-REGN DiscovEHR collaboration, Regeneron Genetics Center, Alan R. Shuldiner, Suganthi Balasubramanian, Goncalo R. Abecasis, Hyun M. Kang, Jonathan Marchini, Eli A. Stahl, Eric Jorgenson, Robert Sanchez, Wolfgang Liedtke, Matthew Anderson, Michael Cantor, David Lederer, Aris Baras, Giovanni Coppola

**Affiliations:** Regeneron Genetics Center, Tarrytown, New York, USA; Regeneron Pharmaceuticals, Inc., Tarrytown, New York, USA; Clinical Trial Service Unit & Epidemiological Studies Unit, Nuffield Department of Population Health, University of Oxford, Oxford, UK; MRC Population Health Research Unit, Nuffield Department of Population Health, University of Oxford, Oxford, UK; Experimental Research Unit from the Faculty of Medicine (UIME), National Autonomous University of Mexico (UNAM)

## Abstract

Human genetic studies of smoking behavior have been so far largely limited to common variations. Studying rare coding variants has potential to identify new drug targets and refine our understanding of the mechanisms of known targets. We performed an exome-wide association study (ExWAS) of smoking phenotypes in up to 749,459 individuals across multiple ancestries and discovered a protective association signal in *CHRNB2* that encodes the β2 subunit of α4β2 nicotine acetylcholine receptor (nAChR). Rare predicted loss-of-function (pLOF) and likely deleterious missense variants in *CHRNB2* in aggregate were associated with a 35% decreased odds for smoking more than 10 cigarettes per day (OR=0.65, CI=0.56-0.76, P=1.9e-8). An independent common variant association in the protective direction (rs2072659; OR=0.96; CI=0.94-0.98; P=5.3e-6) was also evident, suggesting an allelic series. The protective effects of both rare and common variants were detectable to some extent on phenotypes downstream of smoking including lung function, emphysema, chronic obstructive pulmonary disease (COPD) and lung cancer. α4β2 is the predominant nAChR in human brain and is one of the targets of varenicline, a partial nAChR agonist/antagonist used to aid smoking cessation. Our findings in humans align with decades-old experimental observations in mice that β2 loss abolishes nicotine mediated neuronal responses and attenuates nicotine self-administration. Our genetic discovery will inspire future drug designs targeting *CHRNB2* in the brain for the treatment of nicotine addiction.

## Main

Tobacco smoking is one of the greatest hazards to human health globally, accounting for over 200 million disability adjusted life years (DALYs) and 7 million deaths each year^1^. The currently available first line smoking cessation drugs—varenicline and bupropion—were introduced more than two decades ago, even before the human genome project was completed and the genomic revolution started^2–4^. Despite their proven efficacy and wide usage^5^, smoking remains a global health hazard, warranting advancements in smoking related drug discovery efforts that make use of recent innovations in therapeutic design and delivery^6^.

Large scale rare variant association studies have the potential to advance drug discovery^7–10^. Drug designs inspired by naturally occurring genetic variants that protect humans against diseases have been successful in the past, for example, *PCSK9* inhibitors for the treatment of hypercholesterolemia^11–13^. Smoking behavior is strongly influenced by genetics, with twin studies estimating its heritability up to 45%^14^. Both common and rare variants contribute to this high heritability. However, human genetic studies of smoking behavior have so far focused mainly on common variants (those observed in more than 1% of the population)^15–17^. Such genome-wide association studies (GWAS) were successful in identifying genomic regions associated with smoking. In contrast to GWAS, only a very few rare variant studies of smoking exist to date^18,19^. Although such studies have demonstrated that rare variants contribute substantially to smoking heritability, no genes have been confidently linked to smoking based on rare variant associations so far^18,19^.

Unlike common variant associations, rare coding variant associations often pinpoint causal genes^20^, inform effect direction^20,21^, guide follow up experiments^22^ and provide an estimate of the therapeutic efficacy^11,23^ and safety^24^ of targeting a gene or its product. Even for known drug targets, discovering human genetic evidence is valuable as it can improve our understanding of the drug mechanisms and help develop new therapeutic modalities to treat diseases^25^. Hence, with the goal of discovering drug targets for smoking, we undertook a large-scale, exome-wide association study (ExWAS) of smoking behavior involving up to 749,459 individuals. We studied the associations of rare coding variants in the human genome, captured via exome sequencing, with six major smoking phenotypes and a range of secondary phenotypes including diseases in which smoking has been well established as a major risk factor. We also selectively explored the rare variant associations with genes whose associations with smoking via common variants has been realized since the candidate gene era^26^, well replicated by GWAS^15–17,27^ and the mechanisms through which they influence smoking behavior are understood^28,29^. Since we had information on common variants genome-wide for the study participants, we also conducted ancestry-specific and cross-ancestry GWAS meta-analyses for the six smoking phenotypes to validate known loci and to identify novel loci. Finally, we studied the combined influences of both common and rare variants on smoking behavior.

### Exome-wide significant associations

The overall study design is shown in Supplementary Fig. 1. We performed ExWAS meta-analysis for six primary phenotypes—ever smoker, heavy smoker, former smoker, nicotine dependence, cigarettes smoked per day (cig-per-day), and age started smoking—in sample sizes ranging from 112,670 (cig-per-day) to 749,459 (ever smoker). The study cohorts and phenotype definitions are described in Methods, and the cohort-specific sample sizes and participant demographics are summarized in Supplementary Tables 1 and 2. We focused on coding variants of two functional categories: missense variants and predicted loss-of-function (pLOF) variants (frameshift, splice donor, splice acceptor, stop lost, and stop gain) with minor allele frequency (MAF) <0.01. In addition to variant-level associations, we also studied gene-level associations, using burden tests in which either pLOF variants only, or pLOF and likely deleterious missense variants (i.e. predicted to be deleterious by five different algorithms) in a gene are aggregated to create burden masks (or variant sets), which are then tested for association with the phenotypes (Methods)^20^. The burden masks were created using variants at five minor allele frequency (MAF) thresholds (<0.01, <0.001, <0.0001, <0.00001 and singletons) (Supplementary Table 3).

Altogether, we performed 8,417,987 association tests across six smoking phenotypes. Applying a false detection rate (FDR) of 1% (corresponding P-value=4.5e-8), we identified 35 significant associations implicating three genes: *ASXL1, DNMT3A*, and *CHRNB2* (Fig. 1a; Supplementary Fig. 2 and Supplementary Table 4). Although these results were based on analyses where individuals of all ancestries were pooled together (ALL), we found that the results were highly similar to those from a cross-ancestry meta-analysis or a meta-analysis involving only individuals of European ancestry (EUR), suggesting that the results were not influenced by population stratification (Supplementary Fig. 3)

**Figure 1.**
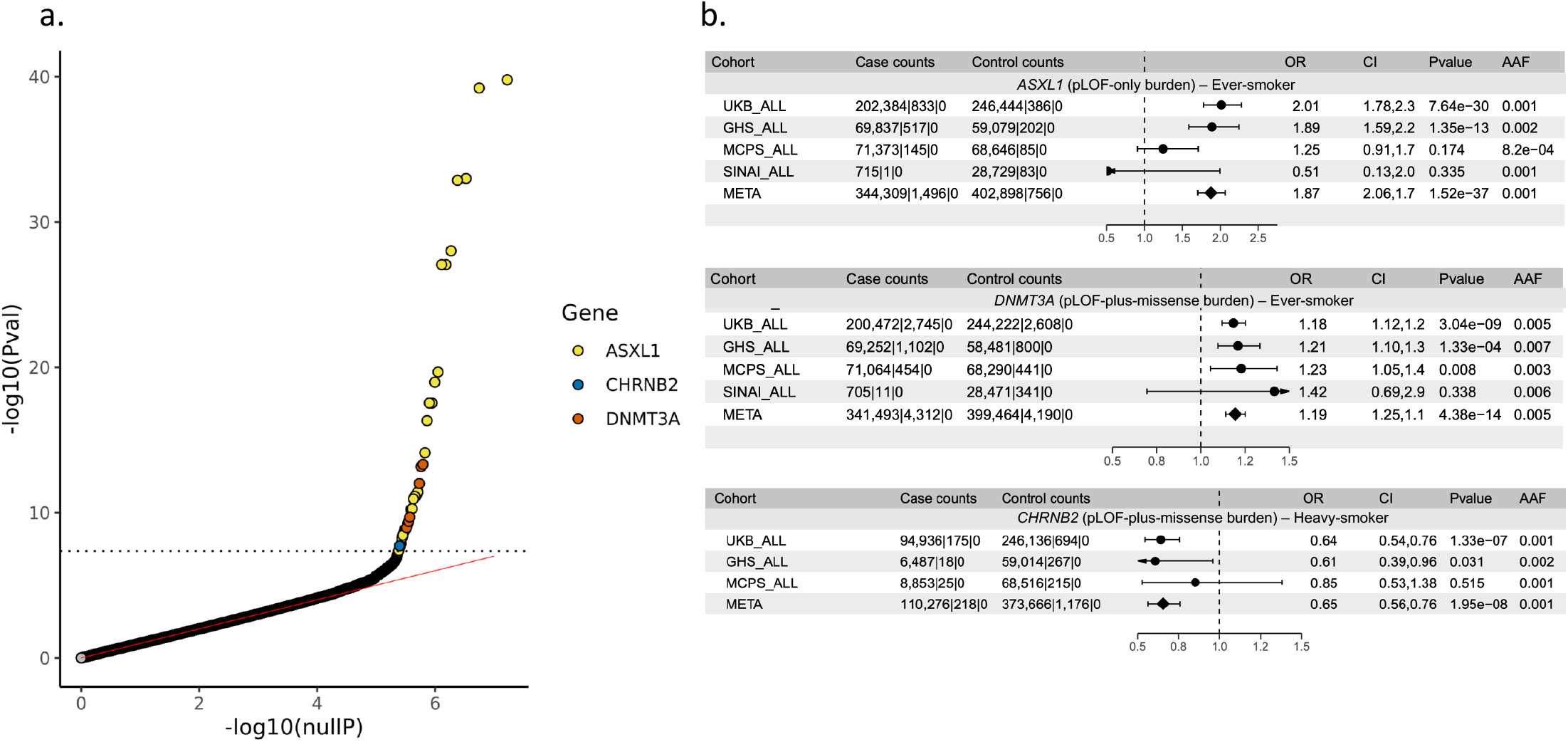
Discovery of rare variants associated with smoking phenotypes. **a**. QQ plot of the rare variant associations with six smoking phenotypes (*ever-smoker, heavy-smoker, former-smoker, nicotine-dependence, cig-per-day* and *age-started-smoking*). The dotted line corresponds to the exome-wide significant threshold, 4.5e-8, defined based on FDR 1% across all the associations combined. **b**. Forest plots of the top burden-trait pairs. For burden mask definitions, refer to Supplementary Table 2. ALL – all ancestries analyzed together; AAF – alternate allele frequency (combined across all variants included in the burden mask)

### Associations of rare and common variants in *CHRNB2*

The primary phenotype that discovered the *CHRNB2* association was *heavy-smoker*, where cases were individuals who smoked at least 10 cigarettes per day either currently or formerly (n=110,494), and controls were individuals who have never smoked in their lifetime (n =374,842). The strongest association was observed for pLOF-plus-missense burden (an aggregate of pLOF and likely deleterious missense variants in *CHRNB2* with MAF< 0.001) where the odds of being a heavy smoker were significantly lower in carriers than non-carriers (OR=0.65; CI=0.56-0.76; P=1.9e-8). The rare variant burden association was independent of any nearby common variant associations with P<0.001 (Supplementary Fig 4; Methods), and the effect estimates were consistently in the protective direction across the three cohorts that contributed to the meta-analysis (Fig. 1b). The protective association was also seen for *ever-smoker* (where individuals who ever smoked regularly in their lifetime were defined as cases, n=345,805) but was less significant compared to heavy smoker, despite a relatively larger sample size, highlighting the importance of phenotype specificity in gene discovery (Supplementary Fig. 5). However, when considering pLOF-only burden (an aggregate of pLOF variants in *CHRNB2* with MAF <0.001)—which provides the strongest evidence on the direction of the association—the association reached at least a nominal level of significance (P<0.05) only for *ever-smoker* but not for *heavy-smoker*, likely because *ever-smoker* captured more pLOF carriers (247 carriers) than *heavy-smoker* (174 carriers), suggesting that a larger sample size at the expense of phenotype specificity is also valuable, particularly at the rarer end of the allele frequency spectrum.

We next studied the association of *CHRNB2* pLOF-plus-missense burden with a range of secondary smoking phenotypes, mainly derived from the UK Biobank (UKB)^30^ participants’ response to lifestyle questionnaire related to smoking (Methods). We also studied the burden associations with a curated list of binary and quantitative health phenotypes related to smoking. The overall association pattern was in line with our main finding that rare pLOF and likely deleterious missense variants in *CHRNB2* in aggregate confer protection against smoking addiction (Supplementary Fig. 6 and Supplementary Table 5). To highlight a few, compared to non-carriers, burden carriers (i.e. carriers of any of the variants included in the pLOF-plus-missense burden mask) showed a decreased risk for emphysema (OR=0.45; CI=0.28-0.71; P=6.9e-4), had better lung function (forced expiratory volume (FEV1): beta=0.05 SD ; CI=0.01-0.09; P=0.01), smoked fewer cigarettes (association seen only in current smokers; beta=-0.23 SD [∼2.3 cigarettes]; CI=-0.45-0; P=0.04), more likely to reduce smoking as a health precaution (OR=2.6; CI=1.0-6.7; P=0.04), more likely to find it fairly easy to not smoke for one day (OR=2.0; CI=1.2-3.4; P=0.007), more likely to wait for 30 to 60 min before smoking the first morning cigarette (OR=1.7; CI=1.0-2.7; P=0.03) and more likely to quit smoking for at least 6 months (OR=1.5; CI=1.1-1.9; P=0.006).

No individual pLOF or missense variants in *CHRNB2* surpassed the study-wide significance threshold, suggesting that our sample sizes were still underpowered to capture single-variant associations. To examine the relative contribution of individual rare variants to the pLOF-plus-missense burden association, we performed gene burden tests iteratively, leaving one variant out of the burden mask each time (leave-one-variant-out (LOVO) analysis^31^). Variants that contribute substantially to the burden association will cause a large drop in the statistical significance when left out. Therefore, such an approach can isolate variants that are mainly driving the association and can help evaluate if a burden association is driven by multiple variants or just a single variant; this is important as in the latter, the inferred effect direction cannot be attributed to all the variants that were included in the burden mask. The LOVO analysis revealed one *CHRNB2* missense variant (rs202079239, Arg460Gly) that contributed the most to the pLOF plus missense burden association as reflected by a substantial drop in the statistical significance and burden mask carrier frequency when that variant was left out (Fig 2a. and Supplementary Table 6). Importantly, even after excluding Arg460Gly, the burden association was still nominally significant with a protective odds ratio (OR=0.57; CI=0.44-0.73; P=1e-5), suggesting that other variants in the burden mask as well contributed to the association (Supplementary Table 6). And the Arg460Gly variant by itself showed a moderately significant protective association with the heavy smoker phenotype (OR=0.58; CI=0.45-0.74; P=2e-5). We found that this variant has been drifted to higher frequency in Finns (gnomAD^32^ MAF=0.0018) compared to non-Finnish Europeans (gnomAD MAF=0.00038; Fig. 2b). Statistical power increases with MAF hence we expected that the protective association of Arg460Gly with smoking or related phenotypes might be detectable in Finngen^33^, a population-based cohort in Finland, despite its smaller samples size compared to UKB. A selective exploration of Arg460Gly with smoking, substance use, and smoking-related lung disease phenotypes in the publicly available data from the FinnGen research project (freeze v7) revealed a significant enrichment for protective associations (hypergeometric test for enrichment P=0.03; Figs. 2c and d and Supplementary Table 7). At least two phenotypes showed nominally significant (P<0.05) protective associations—substance use disorder (excluding alcohol) (OR=0.39; CI=0.21-0.73; P=0.003) and chronic obstructive pulmonary disease (COPD) (OR=0.69; CI=0.49-0.96; P=0.03). Therefore, by exploiting the natural phenomenon of genetic drift in an isolated population, we were able to validate the protective association of *CHRNB2* with smoking-related phenotypes in an independent cohort.

**Figure 2.**
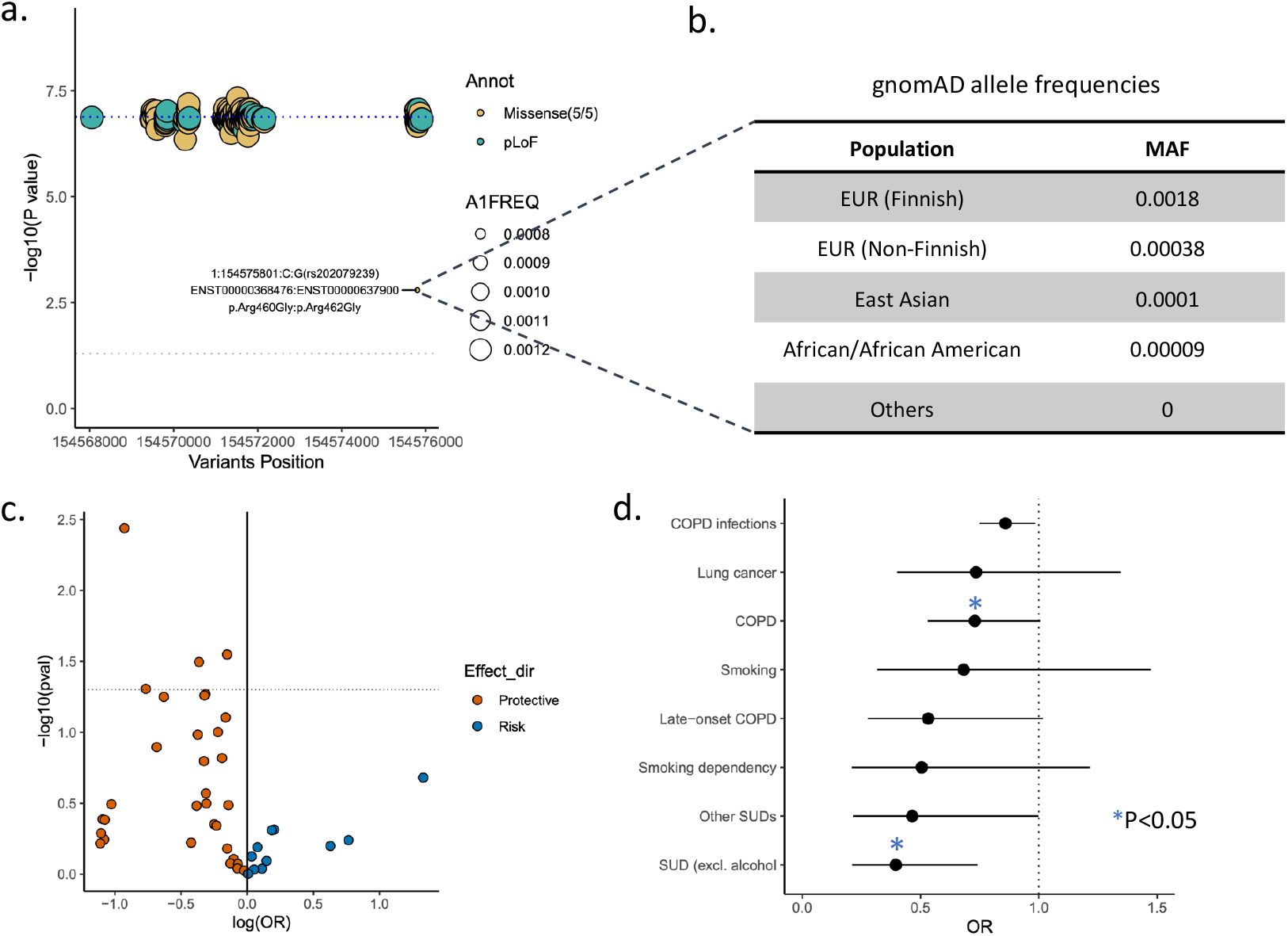
A Finnish enriched missense variant contributes most to the CHRNB2 burden signal. **a**. P values from the leave-one-variant-out (LOVO) analysis of *CHRNB2* rare pLOF-plus-missense burden in the UK Biobank are plotted against the variant positions (hg38). The dotted blue line corresponds to P value of the full burden test. The dotted grey line corresponds to P value=0.05. The variant Arg460Gly that had the largest impact on the LOVO analysis is highlighted and its gnomAD allele frequencies are shown. **c**. Volcano plot showing the PheWAS associations of Arg460Gly related to smoking, substance use, COPD and lung disease endpoints in the Finngen. **d**. OR and 95% confidence intervals of selected phenotype associations of Arg460Gly in the Finngen.

Common-variant associations by themselves often do not pinpoint the causal gene(s); when they do, they mostly bring limited insights on the druggability of the gene. However, when interpreted along with rare coding variant associations, they can offer valuable insights. To this end, we searched for any known common variant GWAS signals near *CHRNB2* that were reported previously for smoking related traits. Liu et al^15^ has reported a GWAS association with cig-per-day near *CHRNB2* where the fine-mapped 95% credible set contained a single variant, rs2072659, located within the 3’ untranslated region (UTR) of *CHRNB2*. This variant showed significant (P<0.05) associations in our dataset with multiple smoking phenotypes, all in the protective direction: *cig-per-day* (Beta=-0.04; CI=-0.02- -0.05; P=7.5e-8), *heavy-smoker* (OR=0.96; CI=0.94-0.98; P=5.3e-6), *ever-smoker* (OR=0.98; CI=0.97-0.99; P=0.001) and *nicotine-dependence* (0.97; CI=0.95-0.99; P=0.003) (Supplementary Fig. 7a and b). In a phenome-wide association study (PheWAS) of this variant across 7,469 phenotypes in two of the largest cohorts (UKB and GHS), the topmost association was with smoking (Supplementary Fig. 7c) In addition, seven out of the top ten associations were with smoking-related phenotypes, all in the protective direction. The findings indicate a convergence between common-variant association with small effect size (OR=0.96) and rare-variant association with large effect size (OR=0.64) hinting a dose-response relationship between natural genetic perturbations of *CHRNB2* and smoking behaviors.

### Associations of CHIP mutations in *ASXL1* and *DNMT3A*

Among the three exome-wide significant genes, *ASXL1* and *DNMT3A* showed strongest associations with most of the smoking phenotypes, including UKB lifestyle questionnaire derived phenotypes and smoking related diseases (Fig 1a and b; Supplementary Fig. 8 and 9; Supplementary Tables 4 and 5). However, both *ASXL1* and *DNMT3A* are known to accumulate somatic mutations in the circulating blood cells with increasing age in the general population, the phenomenon described as clonal hematopoiesis of indeterminate potential (CHIP)^34^. When the DNA source for exome sequencing is peripheral blood, standard exome variant calling workflows capture CHIP mutations along with germline variants, and many studies have exploited this fact to study CHIP mutations using exome sequencing data generated by many large-scale sequencing projects^35,36^. We have previously reported a comprehensive genetic analysis of CHIP where we systematically called somatic variants in participants of the UKB and GHS cohorts and studied their germline associations^36^. It is well known that smoking is strongly associated with CHIP^37,38^, which we have also reported previously^36^. Moreover, the strong association of *ASXL1* CHIP mutations with smoking has emerged in an analysis of UKB’s initial release of ∼50k exomes^38^. Given this background, we were not surprised to learn that the strong associations of *ASXL1* and *DNMT3A* with smoking were fully driven by CHIP mutations. When the associations of *ASXL1* and *DNMT3A* with smoking phenotypes were analyzed using gene burden masks that excluded CHIP mutations, we found associations for neither of the genes with any of the six smoking phenotypes (Fig. 3 and Supplementary Table 8). We further tested the associations of all the eight most recurrently mutated CHIP genes^36^ (*DNMT3A, TET2, ASXL1, PPM1D, TP53, SRSF2, JAK2, SF3B1*) with our six smoking phenotypes in the GHS and UKB, using burden masks created using only CHIP mutations (Supplementary Fig. 10a and Supplementary Table 9). In line with our earlier findings, the strongest associations were seen for *ASXL1* and *DNMT3A*. In addition, we also observed a significant association (P<0.003 based on 1% FDR across eight genes) for *PPM1D* pLOF-only burden with heavy smoker (OR=1.8; CI=1.4-2.3; P=5.5e-7). Notably, certain highly recurrent CHIP driver genes such as *TET2* did not show significant associations with any of the six smoking phenotypes. This suggests that smoking influences the evolution of CHIP mutations through mechanisms that affect not all but only a specific set of genes (Supplementary Fig. 10b). As was previously proposed^38^, it is possible that the chronic inflammation associated with smoking offers a selective advantage to certain CHIP mutation clones to expand, though the precise mechanisms through which smoking influences CHIP mutations are yet to be understood. Also, our findings echo the caution previously raised by many in relation to using exome sequencing data based on blood sample to establish genetic diagnosis for Mendelian diseases in adults^39,40^. In fact, the strongest individual *CHIP* mutation associated with smoking in our analysis was a frame-shift mutation, rs750318549 (p.Gly646fs), in *ASXL1* (*nicotine dependence*: OR=2.41; CI=1.9-3.0; P=7.6e-15), which is pathogenic when it occurs in germline (mostly as de novo), causing a neurodevelopmental disorder called Bohring-Opitz syndrome (Supplementary Table 4)^39^. We found 371 heterozygous carriers in the UKB, who were on average 6.3 yrs. (SE=0.4; P=3.5e-51) older than the non-carriers (average age in carriers=62.9±5.56; non-carriers=56.5±8.1). Observing 371 carriers of a pathogenic variant in a cohort of middle to old aged healthy volunteers strongly points to the somatic origin of this variant.

**Figure 3.**
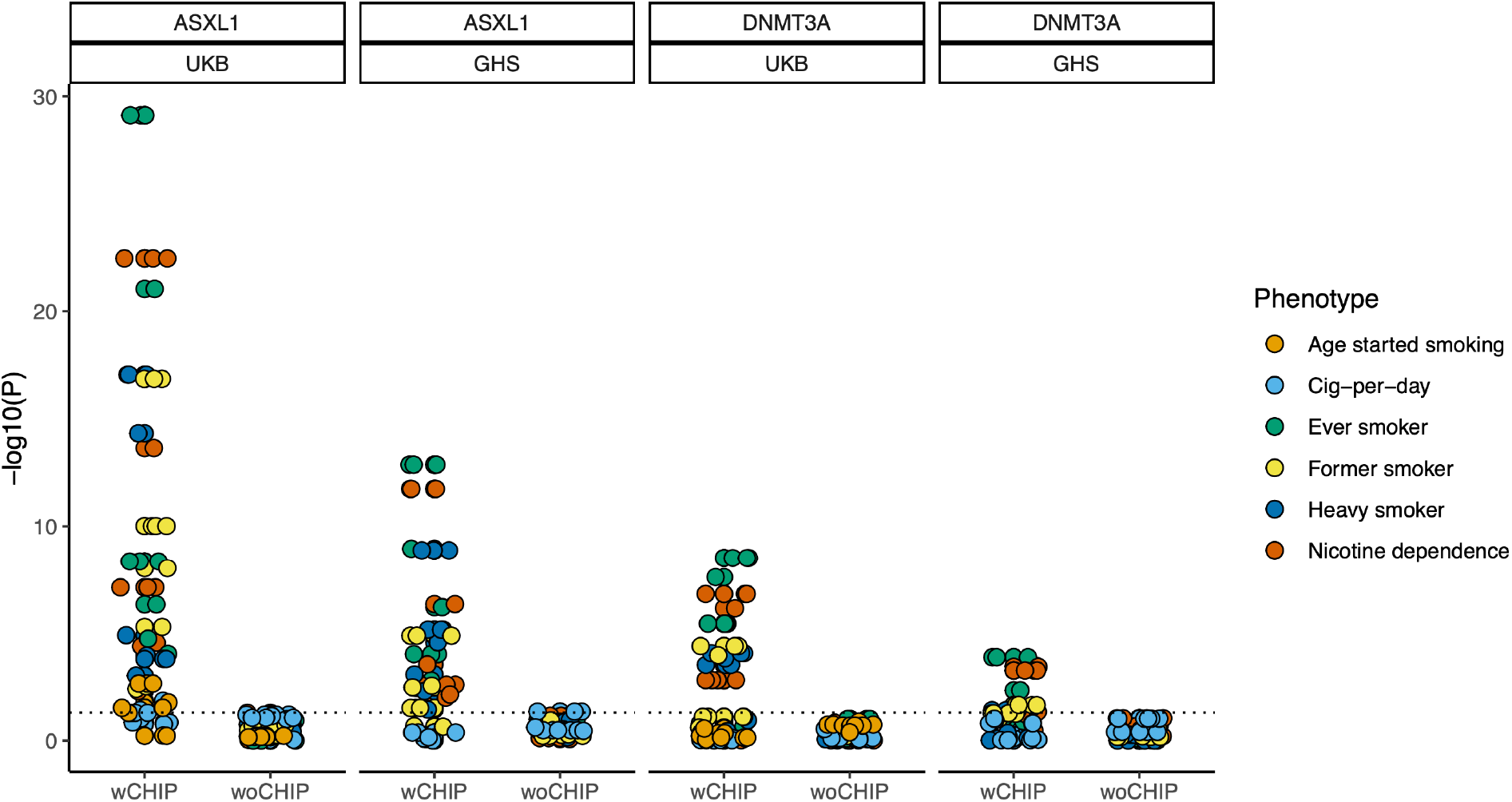
Associations of ASXL1 and DNMT3A based on gene burden masks with and without CHIP variants. We constructed pLOF-only and pLOF plus missense burden masks at five allele frequency thresholds (Methods) using all variants (wCHIP) and variants excluding CHIP variants (wo CHIP). The CHIP variants were identified in the UKB and GHS exome sequencing data using a somatic mutation caller, which we have described in detail previously in Kessler et al 2022. The P values of the burden associations with six primary smoking phenotypes are plotted. Excluding the CHIP variants from the burden masks completely removed all the significant associations suggesting that the ASXL1 and DNMT3A significant burden associations are fully driven by the CHIP variants. CHIP – Clonal Hematopoiesis of Indeterminate Potential

### Association of rare variants in classic smoking genes

Two of the strongest genetic risk loci for smoking that were identified early in the GWAS timeline were locus 15q25.1 containing three nAChR genes (*CHRNA5, CHRNA3* and *CHRNB4*)^28,41^ and 19q13.2 containing a cluster of cytochrome P450 enzyme coding genes (*CYP2A, CYP2B* and *CYP2F* subfamilies), both strongly influence number of cigarettes smoked per day^27,42^. Although none of the genes were exome-wide significant in our analysis, given their strong biological links to smoking, we explored these loci for evidence of any subthreshold rare-variant associations. At the cytochrome P450 locus, we found little evidence for rare variant associations beyond the known common variant signals, even at the subthreshold level of significance (Supplementary Fig. 11 and Supplementary Table 10). But we observed nominal rare variant gene burden associations with *cig-per-day* at 15q25.1 implicating all three nAChRs—*CHRNA5, CHRNA3*, and *CHRNB4*—with effect sizes larger than those observed for common variants. Notably, the largest effect size was observed for the *CHRNB4* pLOF-only rare variant burden where the 13 pLOF carriers smoked on average ∼6.5 cigarettes per day more compared to non-carriers (Beta=0.65 SD; CI=0.14-1.15; P=0.01). This effect size is ∼3 times larger than the largest effect size observed for *CHRNA5* and *CHRNA3* pLOF-only rare variant burden and ∼7 times larger than rs16969968 (∼1 cigarette more; Beta=0.09; CI=0.09-0.10; P=3.8e-125), a well characterized common risk variant at this locus (Supplementary Fig. 11 and Supplementary Table 10). Power calculations based on observed effect sizes suggest that these associations will likely emerge as genome-wide significant when the sample size for ExWAS of cig-per-day reaches between 300k to 500k (Supplementary Fig. 12).

### Cross-ancestry and ancestry-specific associations of common variants

We first performed GWAS for the six primary smoking phenotypes in individuals of European ancestries (EUR) and used these results to analyze SNP based heritability (SNP-h2) and genetic correlations using an EUR ancestry based LD reference panel^43^. Our SNP-h2 estimates were comparable to the estimates reported by GWAS and Sequencing Consortium of Alcohol and Nicotine use (GSCAN) in Liu et al^15^ (Supplementary Fig. 13a; Supplementary Table 11). Also, our GWAS results showed high genetic correlations with the GWAS results from the GSCAN consortium^15^ (excluding UKB) (Supplementary Fig 13b; Supplementary Table 12). The high concordance in the results between our analysis and an analysis in an independent dataset^15^ (GSCAN cohorts excluding UKB) signifies high reproducibility of the polygenic signals of the studied smoking phenotypes. Also, we observed moderate to large genetic correlations across our six phenotypes suggesting that the genetic architecture is shared substantially across the phenotypes (Supplementary Fig. 13c; Supplementary Table 13).

Next, we performed cross-ancestry GWAS meta-analyses for the six primary smoking phenotypes. Across all the phenotypes, in total, we identified 328 linkage disequilibrium (LD) independent loci, of which a majority (94%) are known. This was expected given a GWAS with much larger sample size has been published before^15^ (Supplementary Figs 14a-f; Supplementary Table 15). Among the novel loci, an X chromosome locus we identified for nicotine dependence deserves a special mention as it implicates a nicotinic receptor-related gene. This locus Xq22.1 harbors *TMEM35A* (the closest gene to the index variant), also referred to as *NACHO* (novel acetylcholine receptor chaperone); this gene encodes a molecular chaperone protein that is involved in the assembly of α7, α6β2 and α6β2β3 nAChRs^44^. Mice lacking *TMEM35A* develop hyperalgesia^44^ and we observed that the index variant at this locus is also associated with increased intake of oxycodone, an analgesic medication, in the UKB (OR=1.58; P=0.0001; data from opentargets.org),^45^ suggesting that this locus might influence both smoking and pain phenotypes in humans.

After European ancestries, the second largest proportion (19%) of our study participants were of admixed American ancestries (AMR), mostly from the MCPS cohort^46^. Published GWAS of smoking behavior in AMR ancestries are sparse^47^. In the AMR-specific GWAS, we identified 25 independent loci across the six phenotypes, of which 15 are known and 10 are novel (Supplementary Table 15). The known loci include some of the strongest GWAS signals identified in the EUR-specific GWASs: *CHRNA5*^28^, *CHRNA4*^48^ *and DBH* loci^42^ associated with *heavy-smoker, CYP2A6* locus^27,42^ associated with *former-smoker, NCAM1* locus^49^ associated with *ever-smoker* and *DBH* locus^42^ associated with *heavy-smoker* (Supplementary Table 15). In AMR ancestries, we also identified an X chromosome locus that has been previously linked to smoking in EUR ancestries^17^. Notably, at this locus (with *GPR101* in the vicinity), we identified a genome-wide significant association with *heavy-smoker* in the AMR-specific GWAS (rs1190734; ORAMR=0.83 [0.79-0.88]; PAMR=1.2e-11), but only a nominal association with heavy smoker in the EUR-specific GWAS (OREUR=0.98 [0.97-0.99]; PEUR=0.001). However, the same variant showed genome-wide significant association with cig-per-day in EUR-specific GWAS (BetaEUR=-0.02; PEUR=7.6e-16) corroborating the GWAS signal at this locus reported previously for *cig-per-day*^17^. Whether this locus is associated with cig-per-day in AMR ancestry with a larger effect size compared to EUR is not clear, as we did not have this phenotype in the MCPS cohort at the time of this analysis. Nevertheless, the findings overall suggest that the *GPR101* locus influences smoking behavior in both EUR and AMR ancestries. Regarding the 10 novel loci identified in the AMR ancestries, as expected, many (7 loci) harbored variants that are relatively more common in AMR ancestries than in EUR ancestries, thereby offering higher statistical power for discovery, for example, at 10q21.1, an intergenic locus, we identified a genome-wide significant association with *heavy-smoker* where the index variant is seen in ∼10% of admixed Americans but only ∼0.05% of Europeans; at 8p22 (closest gene: C8orf48), we identified a genome-wide significant association with *ever-smoker* where the index variant is seen in ∼30% of admixed Americans but only ∼7% of Europeans (Supplementary Table 15).

### Interplay between common and rare variants

Large-scale sequencing projects provide increased power to detect additive effects between common and rare variants for many diseases and traits. For example, we have previously demonstrated an additive effect between *GPR75* obesity-protective rare variants and polygenic score (PGS) for obesity based on common variants^10^. We performed a similar analysis to test whether an additive effect is also evident for *CHRNB2* rare variants and smoking PGS. We calculated smoking PGS for the UKB participants based on a GWAS of *ever-smoker* performed in an independent sample (a meta-analysis of our study cohorts and GSCAN leaving out UKB from both)^15^. We binned the UKB individuals into quintiles based on their smoking PGS and quantified the prevalence of heavy smokers in *CHRNB2* pLOF-plus-missense burden mask carriers (the burden mask that showed the strongest association with heavy smoker) and non-carriers. The prevalence of heavy smokers increased in both carriers and non-carriers from lower PGS quintiles to higher, albeit with a small decline from fourth to fifth quintiles (Fig 4; Supplementary Table 16). Importantly, within each of the quintiles, the prevalence of heavy smokers was lower in *CHRNB2* rare variant carriers than non-carriers, demonstrating an additive effect between common and rare variants. The additivity implies that the smoking PGS modifies the penetrance of *CHRNB2* rare variants i.e., smoking behavior varies according to the polygenic background even within rare *CHRNB2* variant carriers.

**Figure 4.**
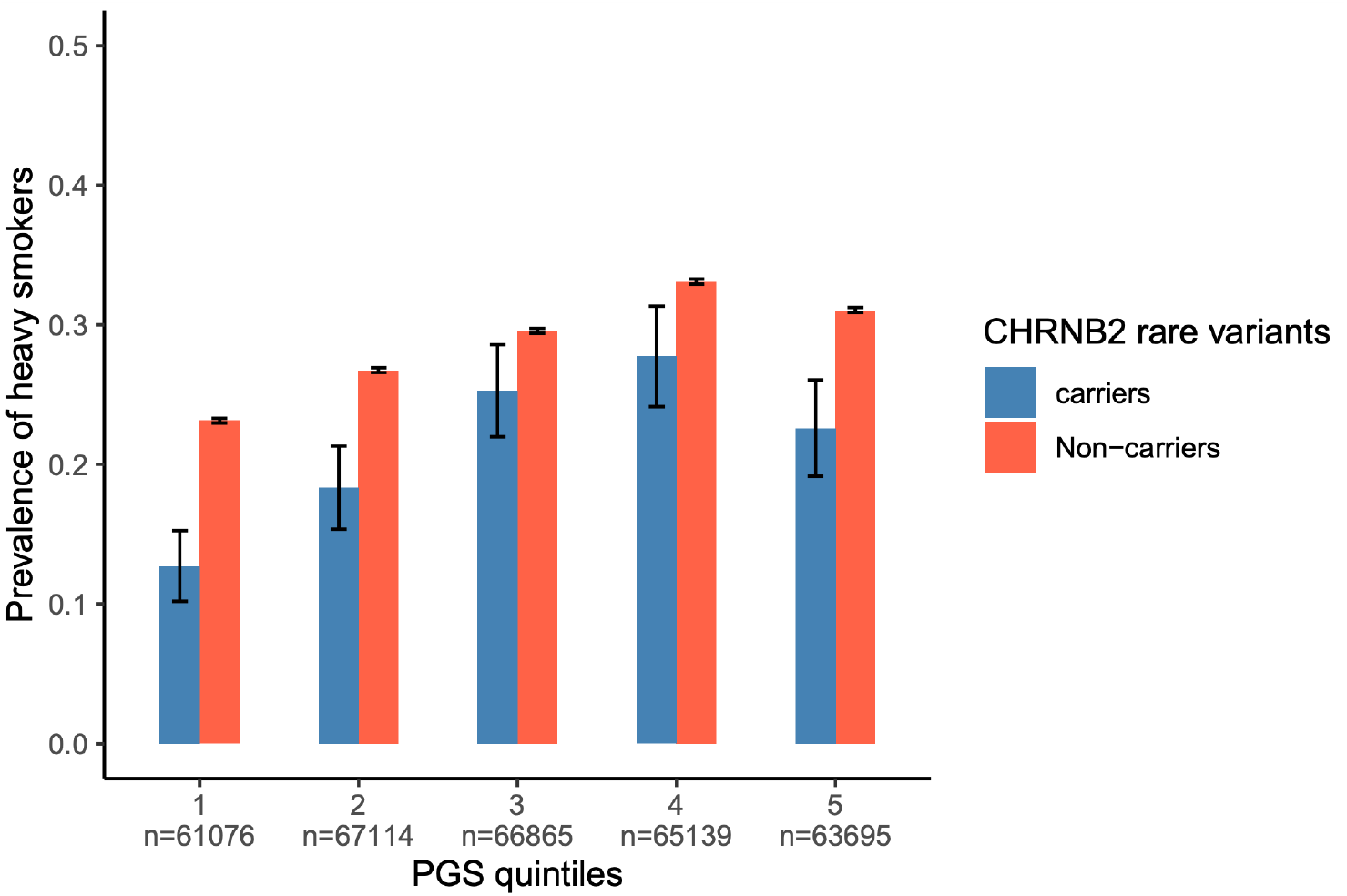
Additive effects between CHRNB2 rare variants and smoking polygenic score. Prevalence estimates heavy smokers in CHRNB2 rare variants carriers and non-carriers within each of the five PGS quintiles in the UK Biobank are plotted. Polygenic score (PGS) was based on a GWAS meta-analysis of ever smoker **(Internal + external without UKB)**. CHRNB2 rare variants mean variants that comprise CHRNB2 pLOF_miss_0.001 burden mask, i.e., pLOF and likely deleterious variants with MAF <0.001. The absolute count of cases and controls in each of the quintiles are reported in the Supplementary Table 16.

## Discussion

GWAS of smoking behavior^15–17^ based on common variants have made tremendous progress in the field with the sample size in the recent GWAS from the GSCAN consortium crossing over 3 million individuals. Such studies have significantly improved our understanding of the polygenic architecture of smoking phenotypes and pointed to genes and pathways underlying smoking behavior, including genes encoding nAChRs, genes involved in nicotine metabolism and dopaminergic and glutamatergic signaling^15^. However, to date, few studies based on whole exome or genome sequencing data have been reported^19,50^ and such studies were conducted in sample sizes insufficient to capture associations at variant and gene level resolutions. Hence, our understanding of the contributions of rare variants to smoking behavior has been minimal so far. To the best of our knowledge, the data we report here represents the largest rare variant study of smoking behavior to date. Our sample sizes were powered to identify associations with a rare variant or an aggregate of rare variants with an odds ratio of 2.5 and above (or 0.4 and below) when present in at least 100 carriers (Supplementary Fig. 15). The fact that our analysis revealed only one germline association indicates that there are no “low hanging fruits” for smoking in the rare variant space other than the *CHRNB2* association that we identified. However, we acknowledge that this interpretation applies only to European populations, and we cannot exclude the possibility there exist rare variants that are more frequent in other ancestries might be discovered in the future in similar or even smaller sample sizes than ours. Nevertheless, we note that 25% of our samples represent non-European ancestries with the largest proportion (19%) representing admixed-Americans. However, the samples sizes, when broken down to individual ancestry groups, are still smaller than what would be necessary to make rare variant discoveries.

The major finding from our analysis is the protective association between *CHRNB2* rare variants and smoking. We show that individuals who carry rare pLOF and likely deleterious missense variants have significantly decreased odds of becoming a heavy smoker. Although the top association was observed for the gene burden that combined both pLOFs and missense variants, the concordant protective effect sizes observed for pLOF-only burden with heavy smoker and ever smoker strengthened our interpretation that what we observe is a loss-of-function and not gain-of-function association. This knowledge is crucial as it directly informs therapeutic hypotheses for drug design. Moreover, we identified a single deleterious missense variant that drifted to higher frequency in Finnish population, which gave us an opportunity to validate the protective associations observed with smoking and smoking-related diseases in the FinnGen study^33^. The finding highlights the value of studying isolated populations with drifted variants to enhance drug target discovery^51^. Another important finding is the convergence of rare and common variant findings of *CHRNB2*. We highlight a common 3’UTR variant that shows protective associations with multiple smoking phenotypes, suggesting that this variant likely decreases *CHRNB2* expression.

Importantly, the odds ratio of the common variant association with heavy smoker was 0.96 as opposed to the odds ratio 0.65 for the pLOF plus missense rare variant burden association with heavy smoker. The pattern reveals a dose-response relationship between the gene and phenotype where varying levels of gene perturbations resulting in proportional effects on the phenotype. We particularly highlight the fact that this variant, though was discovered in an earlier GWAS^15^, did not receive attention as it was buried underneath the hundreds of associations identified by the GWAS, reflecting an important limitation of interpreting common variant findings. However, when interpreted in the light of rare variant findings, that particular common variant association stood out as highly valuable, exemplifying the combined value of GWAS and ExWAS in drug target discovery. Such observations will become frequent in the future with the rapidly growing population-scale ExWASs of human diseases and traits^52^.

*CHRNB2* codes for the β2 subunit of the α4β2 nAChR, which is the predominant nicotinic receptor expressed in the human brain^53^. The role of α4β2 nAChR in mediating nicotine effects has been well characterized by decades of animal studies^54,55^, thanks to the pioneering work of Picciotto and colleagues who first demonstrated in 1995 that deletion of β2 in mice abolished nicotine-mediated effects on avoidance learning and reinforcement behavior^56,57^. However, to the best of our knowledge, what we describe here is the first human genetic evidence supporting the hypothesis that loss of *CHRNB2* protects against nicotine addiction. Importantly, *CHRNB2* can be viewed as a known drug target as it encodes a component of α4β2 nAChR, which, being the major nicotine receptor in the brain, has been the target of most nAChR partial agonists and antagonists developed so far, including cytisine (an α4β2 partial agonist^58^) and varenicline (an α4β2 partial agonist and antagonist^3^). Varenicline is the current drug of choice to aid smoking cessation and was first developed in 1997 by Pfizer based on the molecular structure of cytisine^2,3^. In addition to α4β2, varenicline binds to various other nAChRs in the brain including α7, α3β4 and α6β2. Given the established role α4β2 in mediating the rewarding and reinforcement actions of nicotine, it is believed that the α4β2 antagonistic action of varenicline helps with smoking cessation^3^. Our finding aligns with this hypothesis, emphasizing that human genetics is useful not only to discover new drugs but also to better understand the mechanism of action of old drugs that have been in use for decades, and such knowledge can pave the way for better drug designs with higher efficacy and limited adverse effects.

Limitations of our study include small sample sizes for finer quantitative phenotypes such as *cig-per-day*, which have limited our power to capture associations of genes mediating aversive effects of nicotine (e.g., *CHRNA5*) and those related to nicotine metabolism (e.g., *CYP2A6*)^27,28^. As is often the case, individuals of non-European ancestries were underrepresented in our study cohorts, which has limited the generalizability of the findings to all ancestries^59,60^. However, we involved a substantial number of individuals of admixed American ancestries, who belong to one of the most underrepresented populations in human genetic studies—a step in the right direction. With growing awareness of the importance of diversity in human genetic studies, representation on non-European ancestries is expected to improve in the future studies^59,60^.

To conclude, we have performed a large-scale ExWAS of smoking behavior and identified a protective association between rare coding variants in *CHRNB2* and smoking. The results align with the findings from published knockout animal models and mechanism of action of varenicline that is currently in use to aid smoking cessation and will support future therapeutic developments to treat smoking addiction.

## Methods

### Participating Cohorts

#### UK Biobank

UK Biobank is an open-access, large population cohort of 500,000 individuals established in the UK^30,61^. The participants were, in general, community-dwelling middle aged to old aged volunteers who were recruited between 2006 to 2010 through invitations sent by mail^61^. The participant’ age ranged between 40 to 69 at the time of recruitment. A deep set of phenotypes has been collected from the participants prospectively, including physical, biochemical and multimodal imaging measures, disease history based on electronic health records and a wide range environmental measures obtained via touchscreen and web-based questionnaires. The smoking phenotypes that we studied in this project were based on the information collected through lifestyle and environment touch screen questionnaires (data field category 100058). The health-related phenotypes that we studied including history of lung and vascular diseases are based on ICD-10 codes from the electronic health records or self-reported or a combination of both.

#### GHS

The GHS participants come from Geisinger’s MyCode community health initiative which was established in 2007 to create a bio repository for research projects investigating the molecular and genetic bases of health and disease^62,63^. The participants were patients enrolled in the health care system who consented to participate in the MyCode initiative and gave access to their electronic health records (EHR). The smoking phenotypes that we studied were based on the clinical history of smoking available in the EHR. Finer details on the smoking behavior such as number of cigarettes smoked per day, age started smoking etc. were available for a subset of patients through spirometry questionnaires available in the EHR.

#### MCPS

The MCPS is large prospective cohort of 150,000 individuals recruited between 1998 to 2004 with a major aim to investigate the known and novel risk factors for mortality in individuals of Mexican descent^46,64^. The participants were residents of Coyoacan and Iztapalapa districts of Mexico City. Phenotype data including information on smoking behavior was collected through house-to-house visits through interviewer administered questionnaires.

#### SINAI

The SINAI participants were from the BioMe biobank program of The Charles Bronfman Institute for Personalized Medicine at Mount Sinai Medical Center established in 2007^65^. The BioMe participants are patients enrolled in the Mt Sinai health system, who consented to participate in the BioMe initiative and gave access to their electronic health records. The smoking phenotypes we studied were derived from the EHR.

### Ethical approval and informed consent

All the study participants have provided informed consent and all the participating cohorts have received ethical approval from their respective institutional review board (IRB). The UK Biobank project has received ethical approval from the Northwest Centre for Research Ethics Committee (11/NW/0382)^20,30^. The work described here has been approved by the UKB (application no. 26041)^20^. The GHS project has received ethical approval from the Geisinger Health System Institutional Review Board under project no. 2006-0258^62,63^. The MCPS study has received ethical approval from the Mexican Ministry of Health, the Mexican National Council for Science and Technology, and the University of Oxford^46,64^. The BioMe biobank has received ethical approval from the IRB at the Icahn School of Medicine at Mount Sinai^65^.

### Phenotype definitions

We defined six phenotypes for the primary analysis: a) *ever-smoker*—cases were those who ever smoked regularly (include both former and current smokers) and controls were those who never smoked in their lifetime; b) *heavy-smoker*—cases were those who smoked 10 or more cigarettes per day (include both former and current smokers) and controls were those who never smoked in their lifetime; c) *former-smoker*—cases were those who smoked in the past but not at the present and controls were current smokers; d) *nicotine dependence*—cases were those who had an ICD-10 F17 diagnosis in the EHR and controls were those who did not have an ICD-10 F17 diagnosis; e) *cig-per-day* - number of cigarettes smoked per day in both current and former smokers; f) age started smoking - age when the person first started smoking.

In addition to the six primary phenotypes, we also studied a set of secondary smoking phenotypes primarily derived from the smoking lifestyle questionnaire data in the UKB (data field category 100058). We also studied a selected list of disease phenotypes related to smoking namely lung cancer (ICD-10 C34), COPD (ICD-10 J44), emphysema (ICD-10 J43), chronic bronchitis (ICD-10 J42), peripheral arterial disease (ICD-10 I73), coronary artery disease (ICD-10 I25) and myocardial infarction (ICD-I21).

### Exome sequencing and variant calling

Individuals from all the participating cohorts were exome-sequenced at the RGC. Exome sequencing and variant calling workflows followed in each of the participating cohorts are described in detail elsewhere^10,20,46,62,66^. Briefly, the DNA source for exome sequencing in all the cohorts was peripheral blood. The DNA samples were first enzymatically fragmented into 200 base-pair DNA libraries to which 10 base-pair bar codes were added to facilitate multiplexed operations. Exome regions containing DNA fragments were captured overnight using a modified version of xGen probe from integrated DNA technologies (IDT). The Captured fragments were then PCR amplified and sequenced in a multiplexed way using 75 base-pair paired-end reads on the Illumina NovaSeq 6000 platform. On average 20x coverage was achieved for more than 90% of the target sequences in 99% of the samples.

Sequenced reads are mapped to hg38 reference genome using BWA-MEM to create BAM files. Duplicated reads are marked for exclusion using Picard tool. Then, variant calling was performed at individual sample level using WeCall variant caller to create a per sample gVCF files to enable a sample level filter. Samples with low sequence coverage (<85% of the targeted bases achieving >20x coverage), excess heterozygosity, disagreement between genetic and reported sex, disagreement between exome and array genotype calls and genetic duplicates were removed. The remaining high quality gVCF files are merged into a single project level VCF (pVCF) file using GLnexus joint genotyping tool. A further variant level filter is applied on the multi-sample pVCF file. SNVs with read depth <7 and INDELs with read depth <10 were removed. Also, variants without either at least a single homozygous genotype or a single heterozygous genotype with allele balance ratio >= 0.15 (>=0.20 if INDEL) were removed. The QCed pVCF files are then converted to analysis ready PGEN format using Plink.v2.

### Genotyping and imputation

Genotyping was done using DNA genotyping arrays that varied from cohort to cohort and are reported in detail in cohort specific publications^30,46,62^. Briefly, the UKB participants were genotyped using Applied Biosystems UK BiLEVE Axiom Array or Applied Biosystems UKB Axiom Array, GHS participants, using either the Illumina Infinium OmniExpressExome or Global Screening Array (GSA) and MCPS, SINAI participants, using GSA. Standard quality control procedures were followed to retain only high-quality genotyped variants, which are then used for imputing common variants using TOPMed LD reference panel^67^. For all the cohorts, the imputation was performed in the TOPMed imputation server by uploading the QCed genotypes in randomized batches. Following imputation, we retained only variants with MAF>0.01 and imputation INFO score > 0.8 for the analysis reported in the current study. After all QC, the final number of common variants included in the cross-ancestry meta-analyses ranged from ∼6.7 million for ever smoker to ∼14 million variants for cig-per-day (final number of variants expectedly decreased with increase in the number of cohorts included in the meta-analyses). Appropriate variables for the genotyping arrays and the imputation batches are used as covariates in all the analysis of imputed variants.

### Genetic ancestry inference

Genetic ancestries of the individuals from all the participating cohorts were quantified using a set of common variants that are genotyped directly using the genotyping arrays^20^. We first computed principal components (PCs) in the HapMap3 individuals using the publicly available genotype reference panel^68^; only high confident variants (MAF>0.10, genotype missingness < 5% and Hardy-Weinberg Equilibrium test P > 1e-5) that are common between our dataset and HapMap3 are used for PC calculations. PCs were first computed in the HapMap3 samples on which the rest of the samples are projected. Individuals were assigned to one of five ancestral groups namely Europeans (EUR), African (AFR), admixed Americans (AMR), East Asians (EAS) and South Asians (SAS) if their likelihood for belonging to a particular ancestry > 0.3; the likelihood estimate is calculated using a kernel density estimator (KDE) trained on the HapMap3 PCs^20^.

### Genetic association analysis

Genetic association analyses were done using REGENIE software^31^. REGENIE uses a two-step whole genome regression framework that controls for population stratification and sample relatedness in a cost-effective and computationally efficient way. Briefly, in the step 1, REGENIE computes trait prediction values (also called as local polygenic scores) using a sparse set of genotypes, which are typically the array genotypes. In the step 2, REGENIE computes the variant associations with phenotypes using either logistic or linear regression where the trait prediction values computed in step 1 are included as covariates along with other covariates namely first 20 genetic PCs computed using common variants, first 20 genetic PCs computed using rare variants, age, age squared, sex, interaction term between age and sex and genotyping batches. Specifically, for binary traits with imbalanced case-control ratios, REGENIE uses a fast Firth regression, which has been shown to perform better than saddle point approximation (SPA) correction used in the logistic mixed model approach implemented in software such as SAIGE^69^. For burden analysis, REGENIE first creates a pseudo-genotype, described as burden mask, by collapsing a set of variants (see Supplementary Table 2 for different burden definitions used) into a single categorical variable and then treats this burden mask in the same way as a variant genotype to compute association statistics. For the top burden associations, we performed a sensitivity analysis called leave-one-variant-out (LOVO) implemented in REGENIE. To perform LOVO, REGENIE creates a series of burden masks iteratively for a given set of variants where during each iteration one variant is left out of the burden mask. The created burden masks are then tested for association with the phenotype of interest. For the top burden associations, we also tested if the associations are driven by any nearby common variant signals. For this, we iteratively included the most significant common variant observed within 1Mb on either side of the gene start as covariate in the REGENIE regression analysis until no nearby common variants with P<0.01 were observed. The burden results from the conditional analysis in each of the cohorts were then meta-analyzed together.

### Identification of independent known and novel GWAS loci

To define approximate LD independent GWAS signals, we used conditional and joint analysis (COJO) implemented in the GCTA software^70^. For the LD reference, we used individual level genotype data of randomly sampled 10,000 unrelated individuals of either EUR ancestry (for cross-ancestry and EUR specific GWASs) or AMR ancestry (for AMR specific GWAS). The standard errors of the GWAS summary statistics were adjusted for LD score regression intercept (see section on LD score regression analysis) prior to GCTA-COJO analysis. We defined GWAS loci as “known” if the index variant in the loci is in LD (R2>0.1) with genome-wide significant variants reported previously. LD calculations are done using Plink.v2^71^. Our list of known GWAS loci came primarily from Liu et al 2019^15^. However, before declaring a variant as “novel”, we also manually queried the variants in GWAS catalog to ensure that the variants are not in LD with variants reported in other smoking GWAS publications.

### LD score regression analysis

We calculated SNP-heritability (SNP-h2), that is, the proportion of phenotypic variance explained by the common variants, using LD score regression software^43^. We used an EUR LD reference panel built in-house using a random set of 10,000 unrelated EUR individuals from the UKB following the instructions provided by the authors of the LD score regression software. Genetic correlations were also computed using LD score regression software using the EUR LD reference panel. We used LD score regression also to quantify the population stratification that is known to inflate GWAS association statistics^43^. We computed LD score intercept for all the GWAS runs including the cross ancestry and AMR specific GWASs and then, compared the values to the corresponding GC lambda values. A GC lambda > 1 but an intercept =1 suggests that the observed inflation in the test statistics is fully due to polygenicity. For phenotypes such as smoking that are substantially influenced by environmental factors, it is common to have intercept values slightly above 1 (but still lower than GC Lambda) indicating that there is inflation in test statistics due to factors other than polygenicity e.g., population stratification, cryptic relatedness etc^43^. To remove such inflations, we applied a correction factor^72^ to the test statistics to constrain the LD score intercept close to 1. We scaled the standard errors of the variant associations by a factor of the square root of LD score intercept. This is a better alternative to GC correction (commonly practiced in large-scale consortium GWASs) as GC correction tends to over-correct the statistics removing true polygenic signals^72^. The LD score statistics before and after intercept correction are reported in the Supplementary Table 14. We used EUR LD reference panel even for cross-ancestry as well as AMR specific GWASs as there are no well-established guidelines on how to handle cross-ancestry or admixed ancestry based GWAS results. We acknowledge that this has likely biased the results towards variants that are shared between EUR and other ancestries.

### Polygenic score analysis

We calculated smoking PGS for the UKB participants using SNP weights based on a GWAS of ever smoker conducted in an independent sample. We performed a GWAS meta-analysis of ever smoker across all our cohorts except the UKB. Then, we meta-analyzed the results with the GWAS of ever smoker (without UKB) available from the GSCAN consortium^15^. We then refined the SNP effect sizes in the GWAS summary statistics using PRS-CS software^73^, which uses a Bayesian approach to calculate SNP posterior effect sizes under continuous shrinkage priors based on an external LD reference panel. The refined SNP weights are then used to compute polygenic scores using Plink.v2 software^71^. We then binned UKB individuals into quintiles (five equally sized groups) based on their smoking PGS. Individuals within each quintile are further divided into carriers and non-carriers of *CHRNB2* pLOF or likely deleterious missense variants at MAF<0.001. Prevalence of heavy smokers are then compared between carriers and non-carriers within each quintile. Standard errors of prevalence was calculated using the formula sqrt((Prev - (1-Prev))/N)) where N is number of individuals in the group.

### Power calculations

All power calculations are done in R using the package “genpwr” available from CRAN^74^. In all the cases, we computed effect sizes (beta values) using the function “genpwr.calc” with the following input parameters: power=0.80, calc=“es”, model=“logistic” for binary phenotypes and “linear” for quantitative phenotypes, Alpha=“5e-8” for GWAS and “4.5e-8” for ExWAS, MAF= values ranging from 0 to 0.5, True.model=“additive” and Test.model=“additive”, N=total sample size, case_rate=N cases/N total (for binary phenotypes) and sd_y=1 (for quantitative phenotypes).

## Data Availability

UKB individual-level genotypic and phenotypic data are available to approved investigators via the UK Biobank study (www.ukbiobank.ac.uk/). Additional information about registration for access to the data are available at www.ukbiobank.ac.uk/register-apply/. Data access for approved applications requires a data transfer agreement between the researcher's institution and UK Biobank, the terms of which are available on the UK Biobank website (www.ukbiobank.ac.uk/media/ezrderzw/applicant-mta.pdf). GHS individual-level data are available to qualified academic noncommercial researchers through the portal at https://regeneron.envisionpharma.com/vt_regeneron/ under a data access agreement. The MCPS represents a long-standing collaboration between researchers at the National Autonomous University of Mexico (UNAM) and the University of Oxford. The investigators welcome requests from researchers in Mexico and elsewhere who wish to access MCPS data. If you are interested in obtaining data from the study for research purposes, or in collaborating with MCPS investigators on a specific research proposal, please visit https://www.ctsu.ox.ac.uk/research/prospective-blood-based-study-of-150-000-individuals-in-mexico where you can download the study's Data and Sample Access Policy in English or Spanish. The policy lists the data available for sharing with researchers in Mexico and in other parts of the world. Full details of the data available may also be viewed at https://datashare.ndph.ox.ac.uk/.

## Acknowledgements

We thank the UK Biobank team, their funders, the dedicated professionals from the member institutions who contributed to and supported this work, and the UK Biobank participants. The exome sequencing was funded by the UK Biobank Exome Sequencing Consortium (i.e., Bristol Myers Squibb, Regeneron, Biogen, Takeda, AbbVie, Alnylam, AstraZeneca and Pfizer). This research has been conducted using the UK Biobank Resource under application number 2604.

We thank the MyCode Community Health Initiative participants for taking part in the DiscovEHR collaboration. This research received funding from Regeneron Pharmaceuticals.

We thank the participants of the MCPS cohort. The MCPS has received funding from the Mexican Health Ministry, the National Council of Science and Technology for Mexico, the Wellcome Trust, Cancer Research UK, British Heart Foundation and the UK Medical Research Council. These funding sources had no role in the design, conduct or analysis of the study or the decision to submit the manuscript for publication. Genotyping, exome sequencing and whole genome sequencing was funded through an academic partnership between the National Autonomous University of Mexico, the University of Oxford, Regeneron, AstraZeneca and Abbvie. The computational aspects of this research were supported by the Wellcome Trust Core Award Grant Number 203141/Z/16/Z and the NIHR Oxford BRC. The views expressed are those of the authors and not necessarily those of the NHS, the NIHR or the UK Department of Health.

We thank the participants and investigators of the FinnGen study.

We thank Susan Croll for the helpful discussions on the results of this study.

## Conflicts of interest

The authors declare the following competing interests: Regeneron authors receive salary from and own options and/or stock of the company.

## Notes

### Author Declarations

All the study participants have provided informed consent and all the participating cohorts have received ethical approval from their respective institutional review board (IRB). The UK Biobank project has received ethical approval from the Northwest Centre for Research Ethics Committee (11/NW/0382). The work described here has been approved by the UKB (application no. 26041). The GHS project has received ethical approval from the Geisinger Health System Institutional Review Board under project no. 2006-0258. The MCPS study has received ethical approval from the Mexican Ministry of Health, the Mexican National Council for Science and Technology, and the University of Oxford. The BioMe biobank has received ethical approval from the IRB at the Icahn School of Medicine at Mount Sinai.

