## Supplementary Figures for "Rare coding variants in *CHRNB2* reduce the likelihood of smoking"

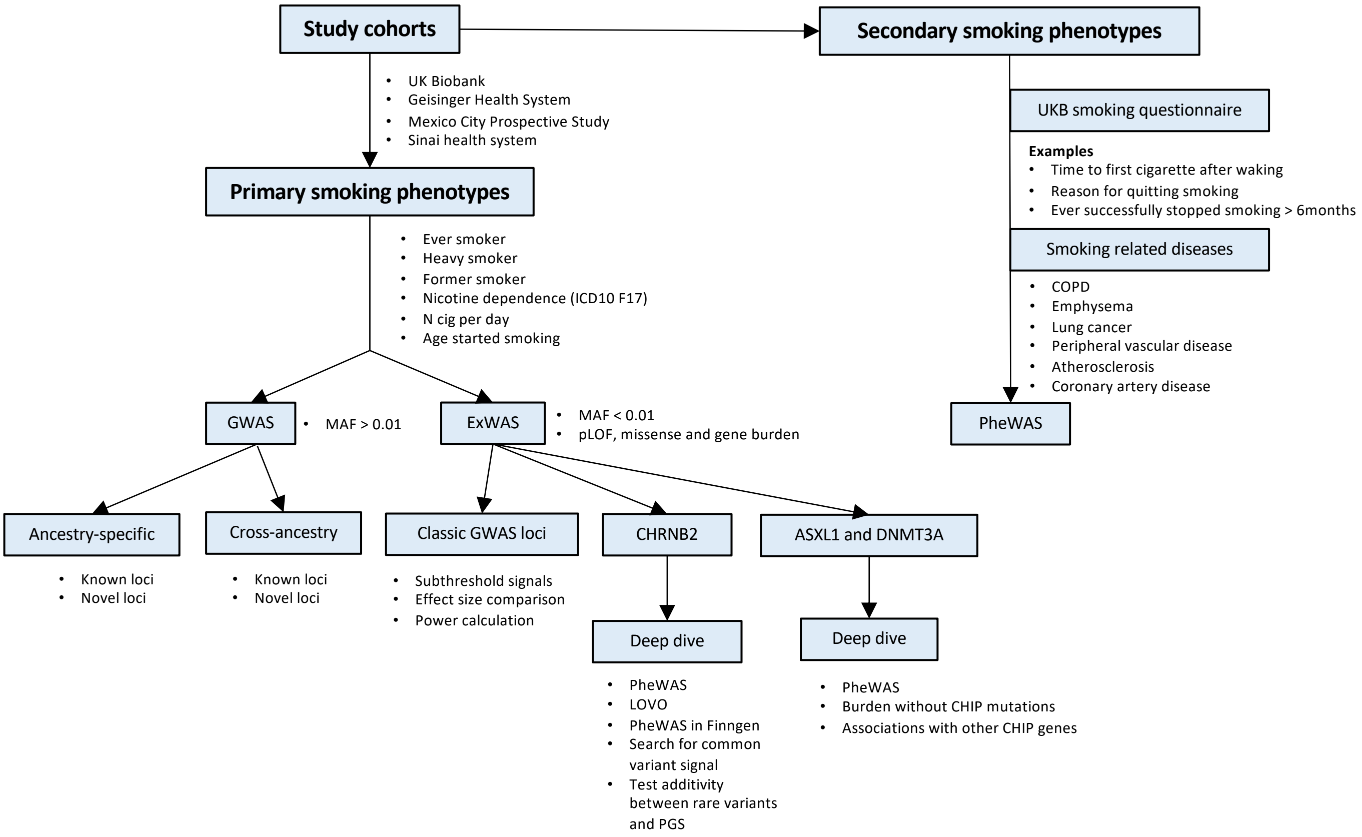


**Supplementary Figure. 1. Overall study design**

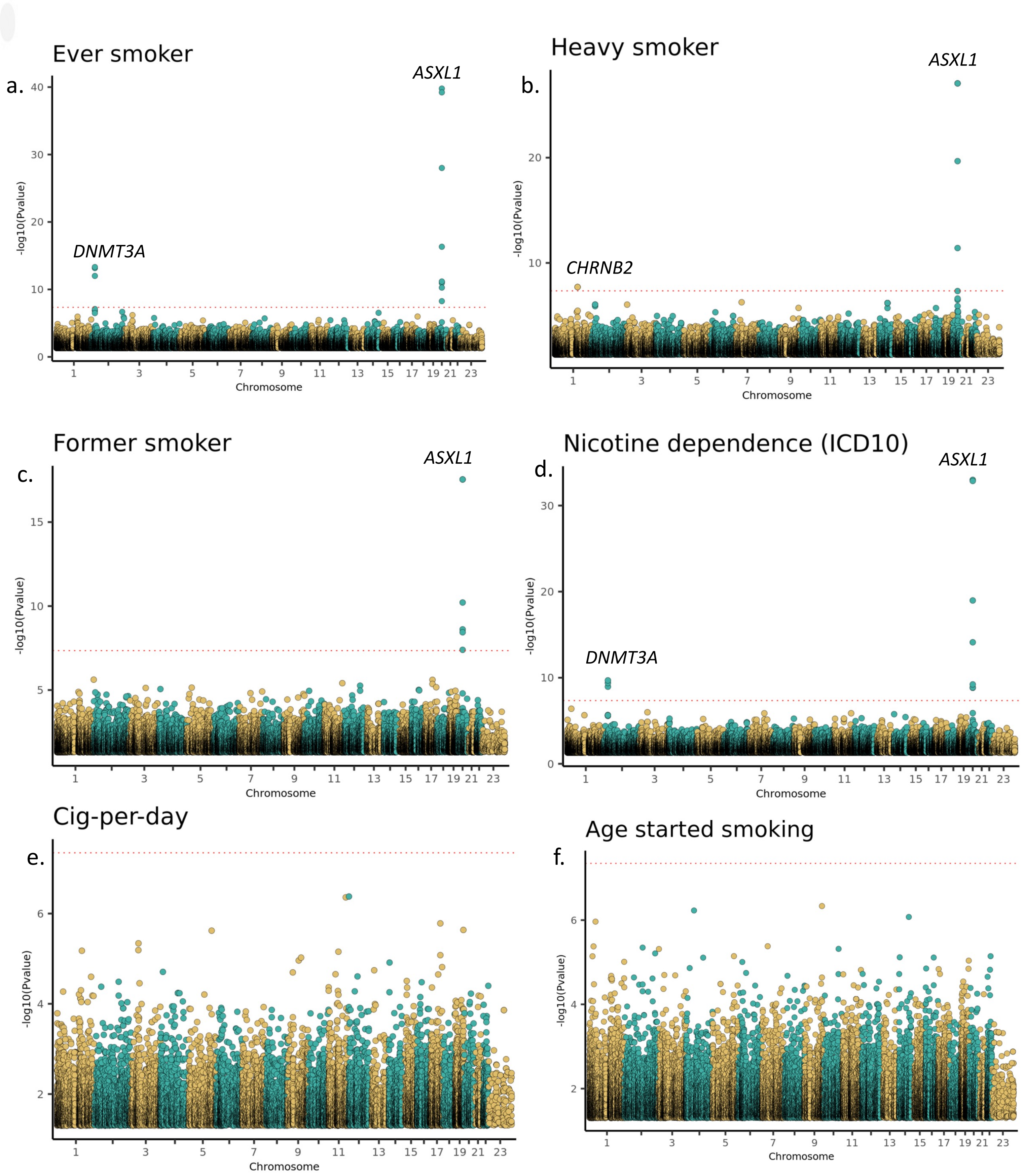


**Supplementary Figure 2. ExWAS Manhattan plots of six smoking phenotypes.** The plots display genome-wide genetic associations of both individual variants (loss of function and missense variants with MAF<0.01) and gene burden masks (pLOF only and pLOF plus likely deleterious missense variants at five MAF cut offs: <0.01, <0.001, <0.0001, <0.00001 and Singletons) with six primary smoking phenotypes (a-f). The exome-wide significance P value threshold was calculated by an applying a false detection rate (FDR) of 1% across the associations of all six smoking phenotypes (8,417,987 association tests in total), which corresponds to P=4.5e-8, marked with a dotted red line in each of the plots.

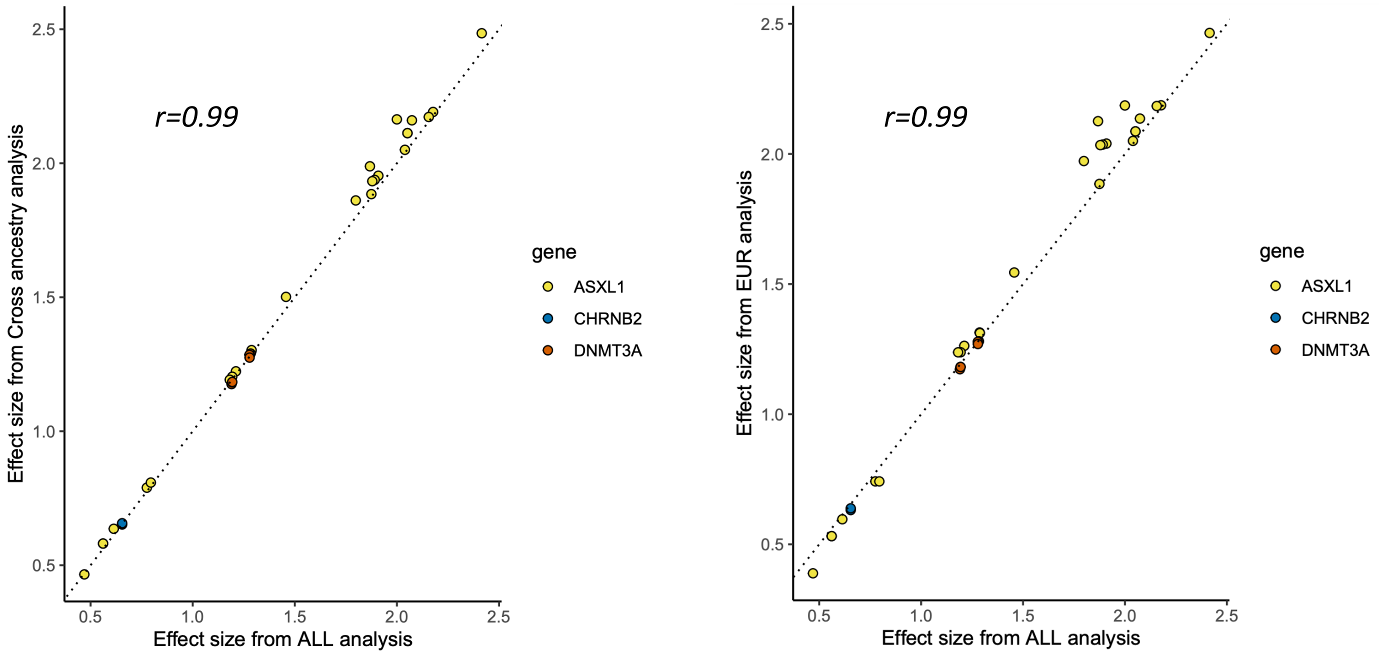


**Supplementary Figure 3. Effect size (odds ratio) comparison of 35 significant ExWAS associations a.** ALL (all ancestries pooled) meta-analysis vs cross ancestry meta-analysis (ancestry specific analysis followed by meta-analysis) and **b.** ALL meta-analysis vs EUR only meta-analysis. Pearson correlation (r) estimates are shown.

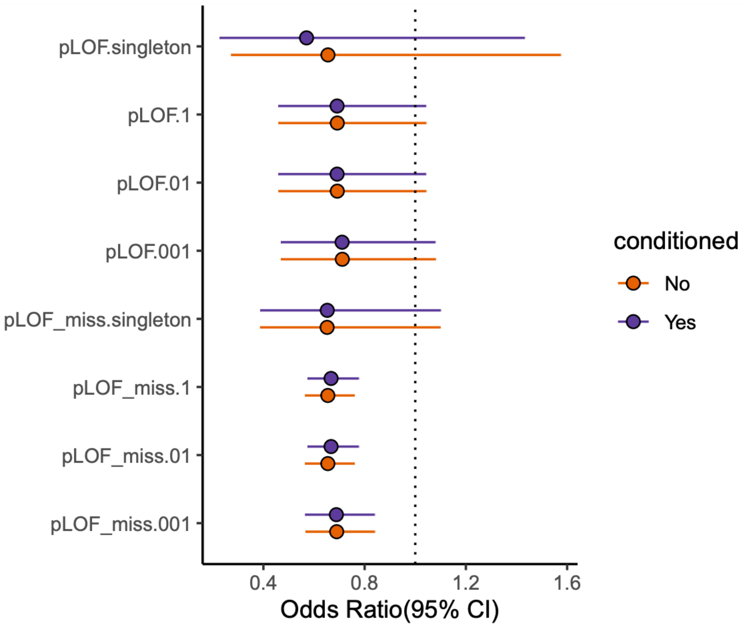


**Supplementary Figure. 4. Analysis of *CHRNB2* burden associations with heavy smoker conditioned on nearby common variants.** CHRNB2 pLOF_miss_0.001 burden association with heavy smoker was computed before and after conditioning on nearby common variants recursively until no variants with 1 Mb on either side of the transcription start site of CHRNB2 has P<0.001. The effect sizes of the burden associations remained the same after conditioning suggesting that the burden associations are independent of any nearby common variant GWAS signals.

### a. ExWAS of heavy smoker

| Cohort | Case counts | Control counts | OR | CI | Pvalue | AAF |
|---|---|---|---|---|---|---|
| CHRNB2_pLOF_miss_0.001 | | | | | | |
| UKB_ALL | 94,936\|175\|0 | 246,136\|694\|0 | 0.64 | 0.54,0.76 | 1.33e−07 | 0.001 |
| GHS_ALL | 6,487\|18\|0 | 59,014\|267\|0 | 0.61 | 0.39,0.96 | 0.031 | 0.002 |
| MCPS_ALL | 8,853\|25\|0 | 68,516\|215\|0 | 0.85 | 0.53,1.38 | 0.515 | 0.001 |
| META | 110,276\|218\|0 | 373,666\|1,176\|0 | 0.65 | 0.56,0.76 | 1.95e−08 | 0.001 |
| CHRNB2_pLOF_0.001 | | | | | | |
| UKB_ALL | 95,091\|20\|0 | 246,755\|75\|0 | 0.66 | 0.41,1.0 | 0.075 | 1.4e−04 |
| MCPS_ALL | 8,874\|4\|0 | 68,684\|47\|0 | 0.76 | 0.26,2.2 | 0.615 | 3.3e−04 |
| GHS_ALL | 6,503\|2\|0 | 59,255\|26\|0 | 1.02 | 0.22,4.8 | 0.982 | 2.1e−04 |
| META | 110,468\|26\|0 | 374,694\|148\|0 | 0.69 | 0.46,1.0 | 0.078 | 1.8e−04 |

### b. ExWAS of ever smoker

| Cohort | Case counts | Control counts | OR | CI | Pvalue | AAF |
|---|---|---|---|---|---|---|
| CHRNB2_pLOF_miss_0.001 | | | | | | |
| UKB_ALL | 202,784\|433\|0 | 246,136\|694\|0 | 0.75 | 0.66,0.85 | 5.33e−06 | 0.001 |
| GHS_ALL | 70,075\|279\|0 | 59,014\|267\|0 | 0.90 | 0.75,1.07 | 0.232 | 0.002 |
| SINAI_ALL | 715\|1\|0 | 28,706\|106\|0 | 0.55 | 0.14,2.13 | 0.383 | 0.001 |
| MCPS_ALL | 71,307\|211\|0 | 68,516\|215\|0 | 0.95 | 0.77,1.18 | 0.673 | 0.001 |
| META | 344,881\|924\|0 | 402,372\|1,282\|0 | 0.82 | 0.90,0.75 | 2.79e−05 | 0.001 |
| CHRNB2_pLOF_0.001 | | | | | | |
| UKB_ALL | 203,183\|34\|0 | 246,755\|75\|0 | 0.51 | 0.34, 0.78 | 0.001 | 1.2e−04 |
| GHS_ALL | 70,332\|22\|0 | 59,255\|26\|0 | 0.73 | 0.40, 1.34 | 0.315 | 1.9e−04 |
| SINAI_ALL | 716\|0\|0 | 28,778\|34\|0 | 0.35 | 0.01,13.55 | 0.570 | 5.8e−04 |
| MCPS_ALL | 71,475\|43\|0 | 68,684\|47\|0 | 0.99 | 0.62, 1.56 | 0.951 | 3.2e−04 |
| META | 345,706\|99\|0 | 403,472\|182\|0 | 0.69 | 0.91, 0.53 | 0.009 | 1.9e−04 |

**Supplementary Figure 5. Forest plots of associations of *CHRNB2* burden masks** with **a.** heavy smoker and **b.** ever smoker. The genotype counts for reference allele homozygous, heterozygous and alternate allele homozygous are shown for cases and controls separately under the columns 'Case counts' and 'Control counts' respectively. UKB – UK Biobank; GHS – Geisinger Health System; MCPS – Mexico City Prospective Study; OR – Odds Ratio; CI – 95% confidence intervals; AAF – Alternate allele frequency;

a.

| Phenotype | Case counts | Control counts | OR | CI | P value | MAF |
|---|---|---|---|---|---|---|
| Why reduced smoking:health precaution | 6,788\|22\|0 | 3,334\|5\|0 | 2.60 | 1.00,6.70 | 0.046 | 0.001 |
| Not smoking for 1 day:fairly easy | 6,696\|25\|0 | 24,230\|46\|0 | 2.00 | 1.20,3.40 | 0.007 | 0.001 |
| First cig after waking in 30−60 mins | 8,588\|28\|0 | 22,242\|43\|0 | 1.70 | 1.00,2.70 | 0.039 | 0.001 |
| Stopped smoking for 6 months | 47,134\|108\|0 | 60,122\|94\|0 | 1.50 | 1.10,1.90 | 0.006 | 9.4e−04 |
| Former smoker − once or twice | 66,328\|216\|0 | 329,768\|777\|0 | 1.40 | 1.20,1.60 | 7.08e−05 | 0.001 |
| Ever smoker | 344,166\|923\|0 | 373,666\|1,176\|0 | 0.82 | 0.75,0.90 | 2.87e−05 | 0.001 |
| Former smoker on most or all days | 108,812\|204\|0 | 287,284\|789\|0 | 0.67 | 0.57,0.78 | 1.96e−07 | 0.001 |
| Heavy smoker | 110,276\|218\|0 | 373,666\|1,176\|0 | 0.65 | 0.56,0.76 | 1.95e−08 | 0.001 |
| Current smoking compared to 10 yrs back:same | 13,552\|22\|0 | 19,811\|54\|0 | 0.63 | 0.40,1.00 | 0.057 | 0.001 |
| Not smoking for 1 day:very difficult | 11,529\|18\|0 | 19,397\|53\|0 | 0.57 | 0.34,0.95 | 0.032 | 0.001 |
| Why reduced smoking: illness | 1,970\|2\|0 | 8,152\|25\|0 | 0.42 | 0.16,1.10 | 0.079 | 0.001 |

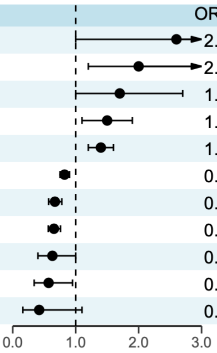


b.

| Phenotype | Case counts | Beta | CI | P value | MAF |
|---|---|---|---|---|---|
| Age started smoking | 33,069\|73\|0 | 0.15 | −0.07,0.37 | 0.183 | 0.001 |
| N unsuccessful smoking quit attempts | 99,411\|187\|0 | 0.11 | −0.02,0.24 | 0.105 | 9.4e−04 |
| FEV1 | 441,463\|1,127\|0 | 0.05 | 0.01,0.09 | 0.012 | 0.001 |
| FVC | 415,708\|1,042\|0 | 0.05 | 0.01,0.09 | 0.015 | 0.001 |
| Age stopped smoking | 108,243\|204\|0 | 0.04 | −0.09,0.17 | 0.541 | 9.4e−04 |
| Age started smoking (former smokers) | 108,261\|204\|0 | 0.03 | −0.10,0.16 | 0.628 | 9.4e−04 |
| Age started smoking | 214,433\|489\|0 | 0.01 | −0.07,0.10 | 0.769 | 0.001 |
| FEV1_FVC | 368,797\|944\|0 | 0.00 | −0.06,0.06 | 0.923 | 0.001 |
| Former smokers−n cig/day | 103,129\|194\|0 | −0.01 | −0.15,0.12 | 0.817 | 9.4e−04 |
| Smoking pack years | 172,837\|374\|0 | −0.02 | −0.11,0.07 | 0.739 | 0.001 |
| Pack years − adult smoking UKB | 133,893\|263\|0 | −0.03 | −0.15,0.09 | 0.603 | 9.8e−04 |
| Cig−per−day | 112,452\|218\|0 | −0.05 | −0.17,0.07 | 0.437 | 9.7e−04 |
| Current smokers−cig/day | 30,741\|71\|0 | −0.23 | −0.45,0.00 | 0.045 | 0.001 |

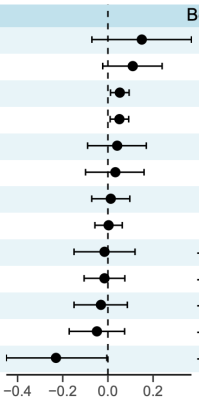


c.

| Phenotype | Case counts | Control counts | OR | CI | P value | MAF |
|---|---|---|---|---|---|---|
| Myocardial Infarction | 44,008\|120\|0 | 378,748\|1,135\|0 | 0.99 | 0.81,1.21 | 0.914 | 0.001 |
| Coronary artery disease | 90,234\|208\|0 | 536,156\|1,336\|0 | 0.92 | 0.79,1.08 | 0.311 | 0.001 |
| Peripheral arterial disease | 18,816\|40\|0 | 325,666\|799\|0 | 0.88 | 0.63,1.22 | 0.430 | 0.001 |
| Lung cancer | 5,142\|11\|0 | 488,834\|1,297\|0 | 0.85 | 0.48,1.50 | 0.575 | 0.001 |
| Family h/o lung cancer | 50,968\|108\|0 | 337,676\|877\|0 | 0.84 | 0.69,1.01 | 0.066 | 0.001 |
| COPD | 34,133\|62\|0 | 315,490\|774\|0 | 0.80 | 0.62,1.03 | 0.082 | 0.001 |
| Chronic bronchitis | 1,413\|2\|0 | 315,490\|774\|0 | 0.77 | 0.27,2.20 | 0.622 | 0.001 |
| Emphysema | 9,695\|13\|0 | 571,428\|1,637\|0 | 0.45 | 0.28,0.71 | 6.92e−04 | 0.001 |

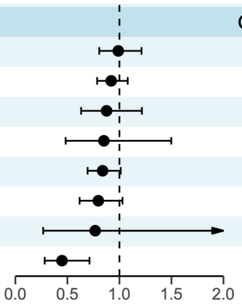


**Supplementary Figure 6. Forest plots of PheWAS associations of CHRNB2 rare pLOF and missense burden** with smoking **a.** binary and **b.** quantitative phenotypes (six primary phenotypes plus lifestyle questionnaire derived secondary phenotypes in the UK Biobank; only associations with P<0.1 are shown) and **c.** smoking related health conditions FEV1 – Forced expiratory volume in 1 sec; FVC – Forced vital capacity; FEV1_FVC – FEV1:FVC ratio; COPD – Chronic obstructive pulmonary disease; OR – Odds Ratio; CI – 95% Confidence Intervals; MAF – minor allele frequency

a.

| rs2072659 | | | | | | |
|---|---|---|---|---|---|---|
| Phenotype | Case counts | Control counts | OR | CI | P value | MAF |
| Former smoker | 152,405\|30,974\|1,666 | 85,763\|13,608\|686 | 1.01 | 0.99,1.03 | 0.475 | 0.086 |
| Ever smoker | 286,174\|56,274\|3,068 | 331,007\|65,493\|3,612 | 0.98 | 0.97,0.99 | 0.001 | 0.091 |
| Nicotine dependence | 52,732\|11,595\|618 | 376,450\|86,170\|4,978 | 0.97 | 0.95,0.99 | 0.003 | 0.102 |
| Heavy smoker | 89,354\|19,684\|1,129 | 307,399\|61,858\|3,467 | 0.96 | 0.94,0.98 | 5.33e−06 | 0.094 |

b.

| rs2072659 | | | | | |
|---|---|---|---|---|---|
| Phenotype | Case counts | Beta | CI | P value | MAF |
| Age started smoking | 180,504\|32,221\|1,722 | 0.00 | −0.01, 0.01 | 0.682 | 0.083 |
| Smoking pack years | 142,014\|29,300\|1,631 | −0.02 | −0.03,−0.01 | 2.78e−04 | 0.094 |
| Cig−per−day | 90,301\|21,008\|1,217 | −0.04 | −0.05,−0.02 | 7.58e−08 | 0.104 |

c.

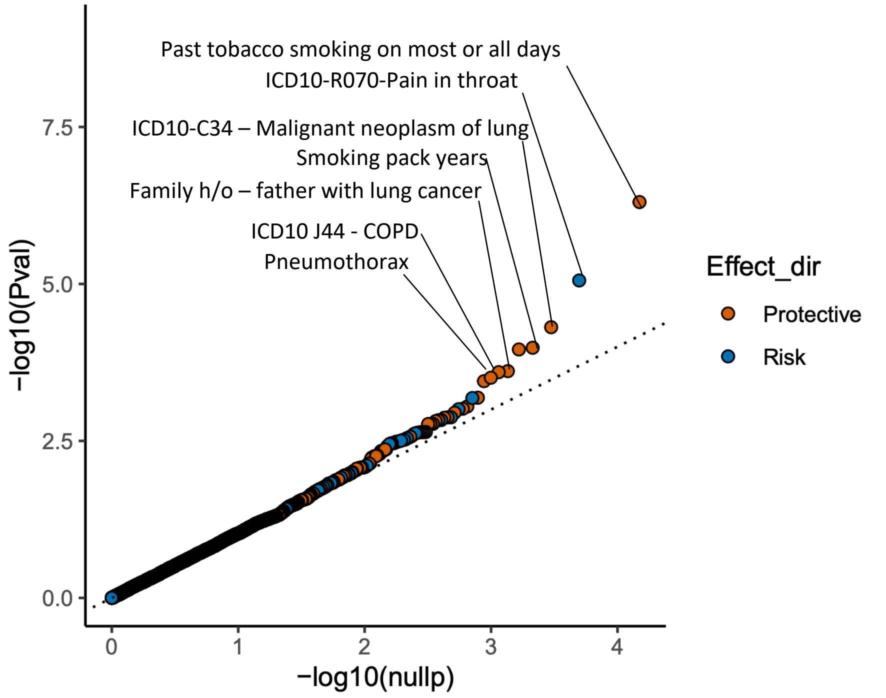


**Supplementary Figure 7. Association of a *CHRNB2* common 3' UTR variant with smoking.** a. forest plots of associations with **a.** binary and **b.** quantitative smoking phenotypes. **c.** QQ plot of PheWAS of the common variant in UKB and GHS cohorts. The top 10 associations are labelled. Protective - OR <1 or beta <0 and risk – OR >1 or beta >0

a.

| Phenotype | Case counts | Control counts | OR | CI | P value | MAF |
|---|---|---|---|---|---|---|
| Current smoker on most or all days | 33,309\|214\|0 | 395,981\|994\|0 | 2.90 | 2.40,3.40 | 1.34e−34 | 0.001 |
| Nicotine dependence | 63,122\|402\|0 | 445,114\|1,342\|0 | 2.18 | 1.92,2.47 | 1.04e−33 | 0.001 |
| Heavy smoker | 109,989\|505\|0 | 374,169\|673\|0 | 2.05 | 1.80,2.33 | 8.74e−28 | 0.001 |
| >1 household member smokes | 4,685\|14\|0 | 392,239\|985\|0 | 1.90 | 1.10,3.40 | 0.029 | 0.001 |
| Ever smoker | 343,594\|1,495\|0 | 374,169\|673\|0 | 1.89 | 1.72,2.08 | 1.67e−40 | 0.001 |
| Why reduced smoking:financial reasons | 3,822\|40\|0 | 6,248\|39\|0 | 1.80 | 1.10,2.80 | 0.010 | 0.003 |
| Former smoker on most or all days | 108,555\|461\|0 | 287,544\|529\|0 | 1.70 | 1.50,1.90 | 4.71e−15 | 0.001 |
| Why stopped smoking:doctor's advice | 7,992\|60\|0 | 78,863\|318\|0 | 1.70 | 1.20,2.30 | 5.98e−04 | 0.002 |
| Light smokers: <=100 smokes/lifetime | 55,300\|156\|0 | 64,028\|100\|0 | 1.50 | 1.20,1.90 | 0.001 | 0.001 |
| Likelihood of resuming smoking:probably no | 10,998\|50\|0 | 95,135\|395\|0 | 1.50 | 1.10,2.00 | 0.019 | 0.002 |
| Former smokers−hand rolled cigarettes | 7,394\|41\|0 | 101,022\|419\|0 | 1.40 | 1.00,2.10 | 0.051 | 0.002 |
| Current smoker−hand rolled cigarettes | 7,910\|64\|0 | 25,354\|150\|0 | 1.40 | 1.00,1.90 | 0.031 | 0.003 |
| Why stopped smoking:illness | 13,263\|73\|0 | 73,592\|305\|0 | 1.30 | 0.96,1.70 | 0.091 | 0.002 |
| Former smoker − once or twice | 66,432\|112\|0 | 329,667\|878\|0 | 0.75 | 0.62,0.91 | 0.003 | 0.001 |
| Current smoker−manufactured cigarettes | 23,091\|124\|0 | 10,173\|90\|0 | 0.75 | 0.56,1.00 | 0.060 | 0.003 |
| Why stopped smoking:health precautions | 68,829\|265\|0 | 18,026\|113\|0 | 0.75 | 0.59,0.94 | 0.012 | 0.002 |
| Likelihood of resuming smoking:definitely no | 94,400\|391\|0 | 11,733\|54\|0 | 0.66 | 0.49,0.90 | 0.008 | 0.002 |
| Why reduced smoking:health precaution | 6,769\|41\|0 | 3,301\|38\|0 | 0.59 | 0.37,0.94 | 0.027 | 0.003 |
| Former smoker | 184,221\|934\|0 | 99,873\|397\|0 | 0.56 | 0.49,0.64 | 2.88e−18 | 0.002 |
| First cig after waking in >2 hrs | 3,594\|9\|0 | 27,120\|178\|0 | 0.52 | 0.28,0.95 | 0.032 | 0.003 |

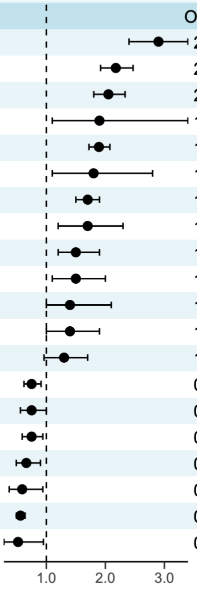


b.

| Phenotype | Case counts | Beta | CI | P value | MAF |
|---|---|---|---|---|---|
| Age stopped smoking | 107,989\|458\|0 | 0.36 | 0.28, 0.45 | 3.35e−16 | 0.002 |
| Pack years − adult smoking UKB | 133,529\|627\|0 | 0.31 | 0.24, 0.39 | 6.27e−16 | 0.002 |
| Smoking pack years | 172,452\|759\|0 | 0.29 | 0.23, 0.36 | 4.29e−18 | 0.002 |
| N unsuccessful smoking quit attempts | 99,179\|419\|0 | 0.12 | 0.03, 0.21 | 0.008 | 0.002 |
| FEV1_FVC | 368,602\|1,139\|0 | 0.08 | 0.02, 0.13 | 0.005 | 0.001 |
| FVC | 415,655\|1,095\|0 | −0.07 | −0.11,−0.03 | 9.03e−04 | 0.001 |
| Age started smoking (former smokers) | 108,007\|458\|0 | −0.09 | −0.18,−0.01 | 0.035 | 0.002 |
| Age started smoking | 214,107\|815\|0 | −0.11 | −0.17,−0.04 | 0.001 | 0.001 |
| FEV1 | 441,264\|1,326\|0 | −0.13 | −0.17,−0.09 | 3.08e−11 | 0.001 |
| Age started smoking | 32,930\|212\|0 | −0.18 | −0.31,−0.04 | 0.008 | 0.003 |

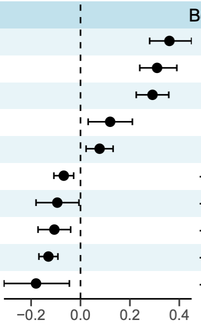


c.

| Phenotype | Case counts | Control counts | OR | CI | P value | MAF |
|---|---|---|---|---|---|---|
| Lung cancer | 5,091\|62\|0 | 489,052\|1,079\|0 | 2.34 | 1.73,3.2 | 3.18e−08 | 0.001 |
| COPD | 33,853\|342\|0 | 315,281\|983\|0 | 1.74 | 1.52,2.0 | 4.48e−15 | 0.001 |
| Emphysema | 9,604\|104\|0 | 571,196\|1,869\|0 | 1.53 | 1.22,1.9 | 2.39e−04 | 0.001 |
| Chronic bronchitis | 1,403\|12\|0 | 315,281\|983\|0 | 1.53 | 0.78,3.0 | 0.219 | 0.001 |
| Peripheral arterial disease | 19,119\|197\|0 | 325,260\|864\|0 | 1.52 | 1.27,1.8 | 6.65e−06 | 0.001 |
| Myocardial Infarction | 44,284\|342\|0 | 378,580\|1,062\|0 | 1.28 | 1.12,1.5 | 3.70e−04 | 0.001 |
| Coronary artery disease | 89,138\|655\|0 | 532,380\|1,436\|0 | 1.22 | 1.10,1.4 | 2.44e−04 | 0.001 |
| Family h/o lung cancer | 50,914\|162\|0 | 337,699\|854\|0 | 1.05 | 0.88,1.2 | 0.568 | 0.001 |

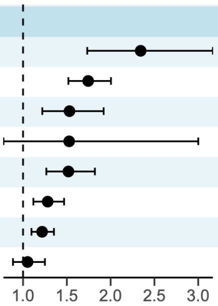


**Supplementary Figure. 8. Forest plots of PheWAS associations of ASXL1 rare pLOF only burden** with smoking **a.** binary and **b.** quantitative phenotypes (primary and secondary-questionnaire derived phenotypes in the UK Biobank; only associations with P<0.1 are shown) and **c.** smoking related health conditions

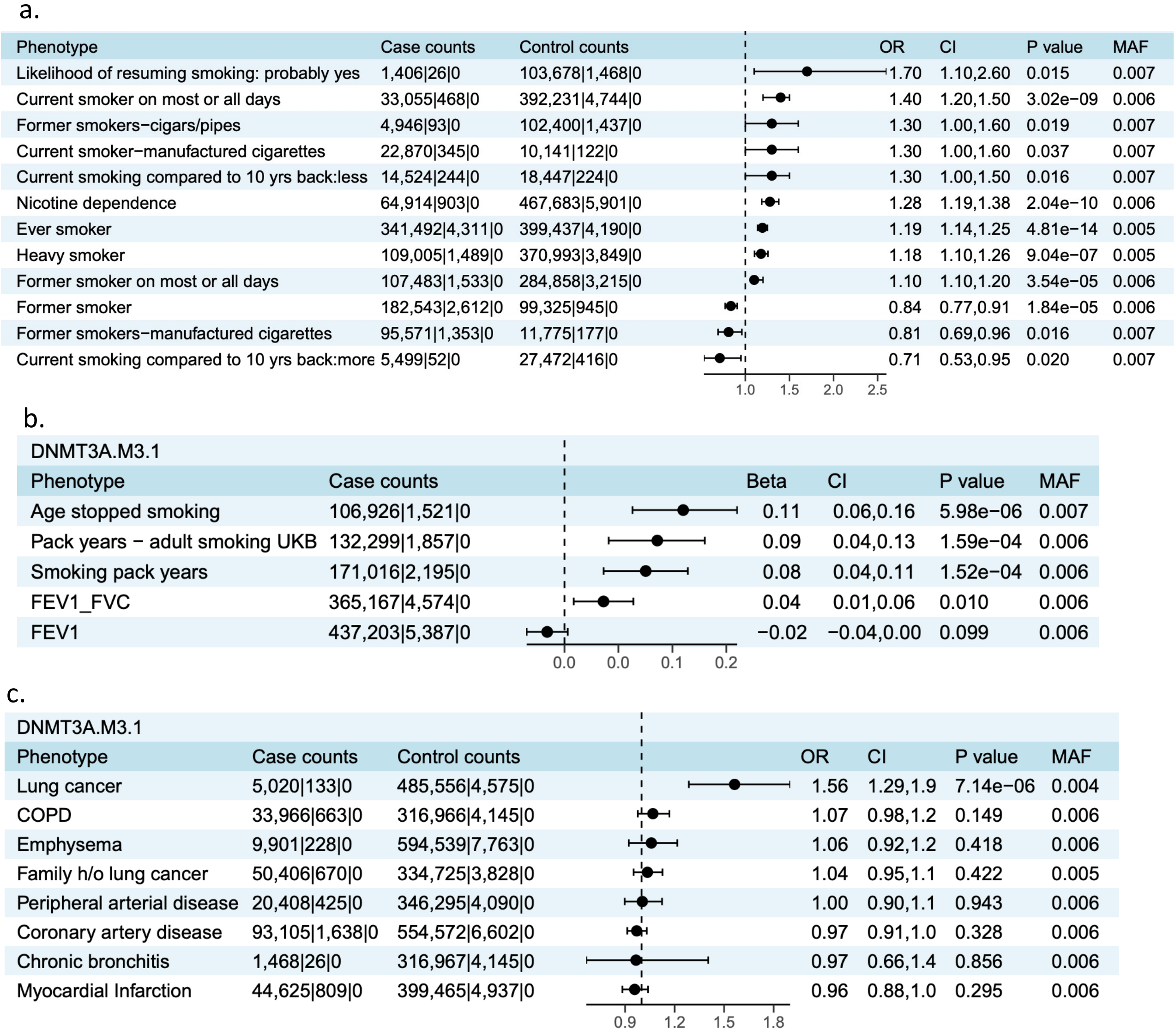


**Supplementary Figure 9. Forest plots of PheWAS associations of DNMT3A rare pLOF plus missense burden** with smoking **a.** binary and **b.** quantitative phenotypes (primary and secondary-questionnaire derived phenotypes in the UK Biobank; only associations with P<0.1 are shown) and **c.** smoking related health conditions

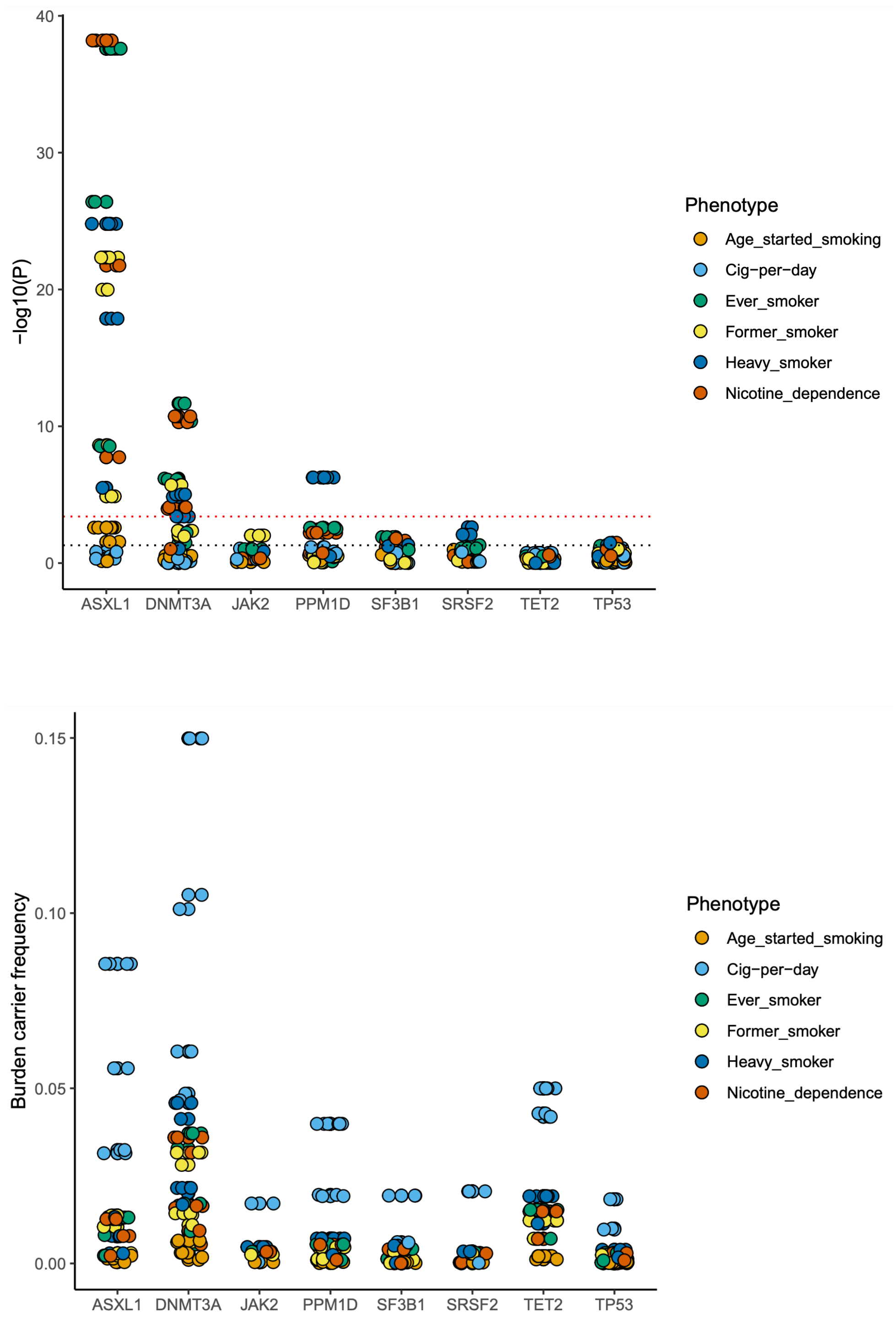


**Supplementary Figure 10. Burden associations of eight most recurrent CHIP genes with smoking.** The burden masks are created by collapsing only CHIP mutations (identified using a somatic mutation caller) under different definitions (Supplementary Table 2) and testing their associations with the six smoking phenotypes. The P values of the associations are plotted in **a.** in –log10 scale. The red dotted line corresponds to a P=0.0003 at FDR 1% threshold applied across all the eight genes. The black dotted line corresponds to P=0.05. The burden allele frequencies are plotted in **b.** to illustrate that some genes such as TET2, despite harboring CHIP mutations in high frequency, did not show significant associations with smoking.

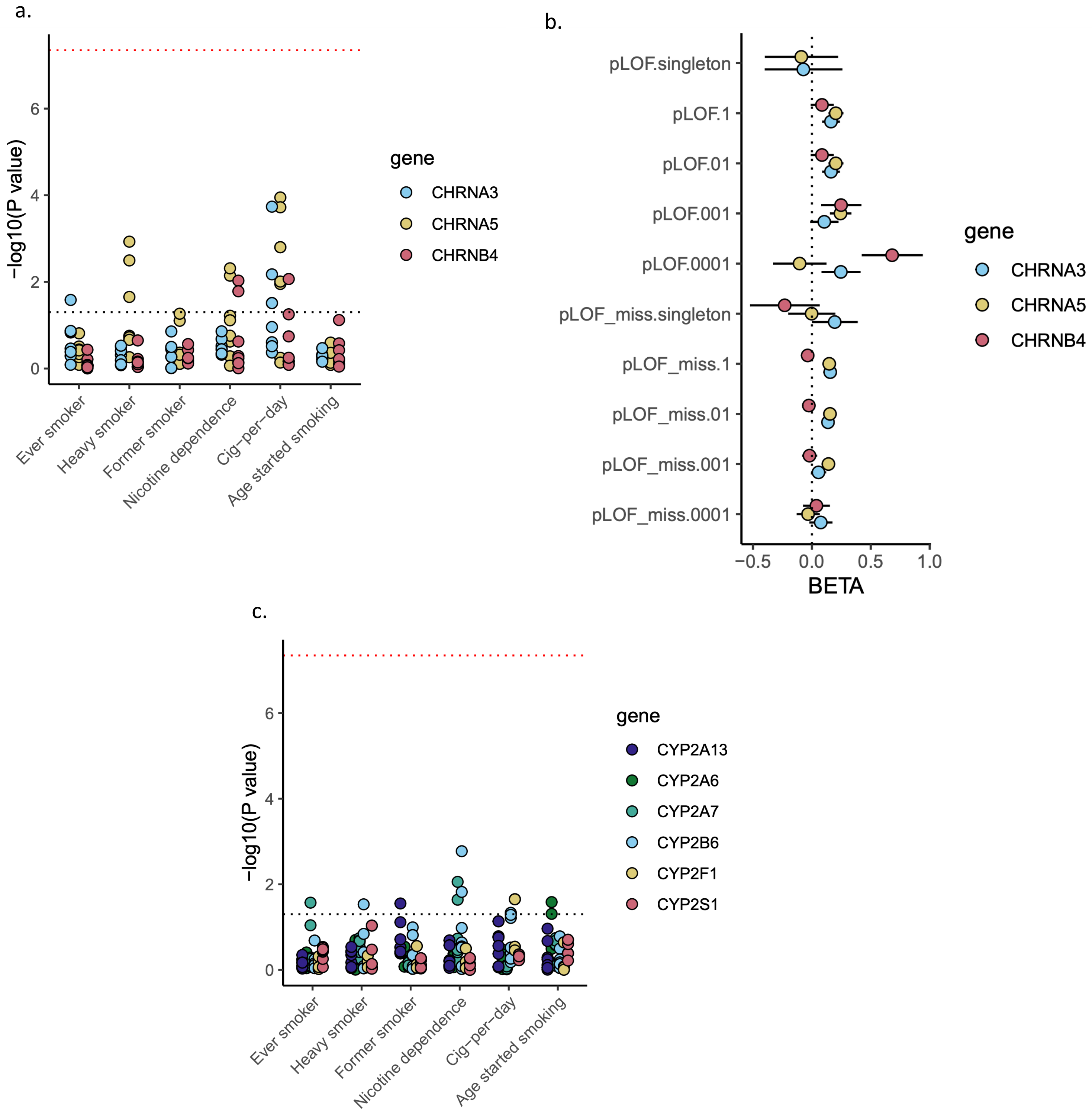


**Supplementary Figure 11. Rare variant associations at the classic GWAS loci. a.** Associations of 10 burden masks (Supplementary Table 2) of *CHRNA5, CHRNA3 and CHRNB4* (at smoking GWAS locus 15q25.1) with six smoking phenotypes are plotted. The red dotted line corresponds to exome-wide significant threshold P value=4.5e-8. The black dotted line corresponds to P=0.05. **b.** the effect sizes (Beta values) and standard errors corresponding to the data points in fig a. are plotted. c. Associations of 10 burden masks of CYP450 family of genes at smoking GWAS locus 19q13.2 are plotted.

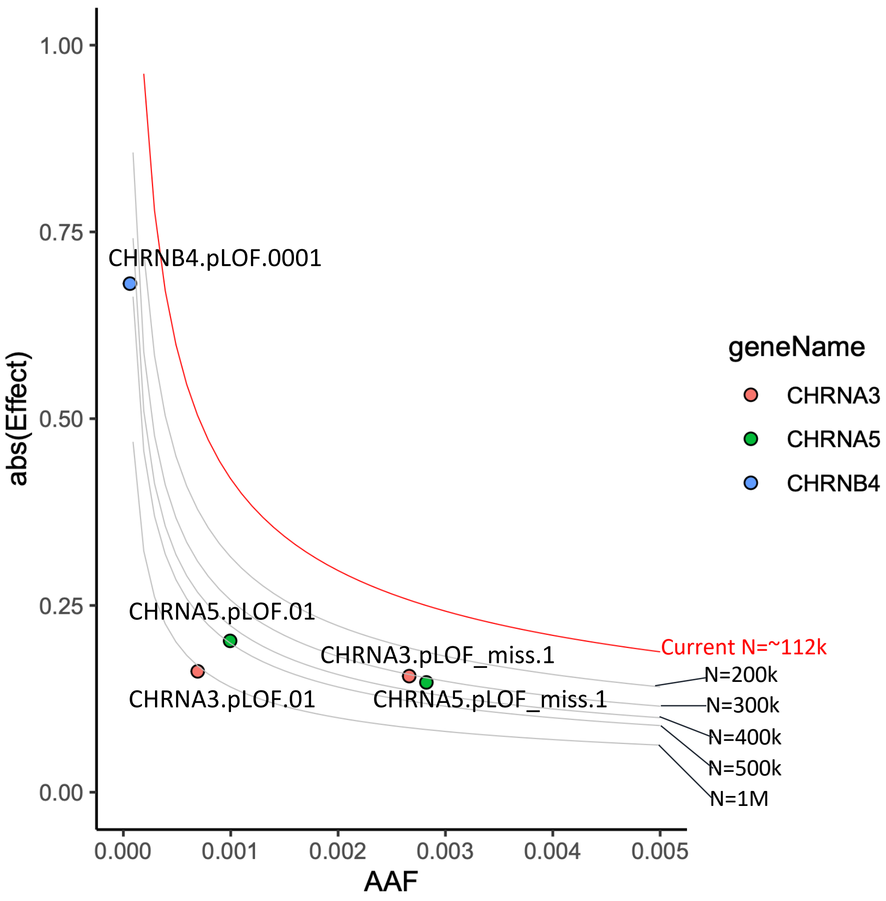


**Supplementary Figure 12. Power calculations for rare variant discovery at the CHRNA5 GWAS locus.** Assuming an 80% power and P value of 5e-8, detectable effect sizes at various minor allele frequency values were calculated for the current sample size of cig-per-day (the smoking trait most associated with CHRNA5 locus) as well for a series of sample sizes up to 1 million. The observed effect sizes for pLOF only burden and pLOF and missense burden associations of CHRNA5, CHRNA3 and CHRNB4 are plotted; all the points lay below the red line, which marks the detection limit of our current sample size, suggesting that we are underpowered. Based on the intersections of the grey lines with the points marking the observed effect sizes, we can approximately guess what sample size will be required to detect these burden signals at P value 5e-8.

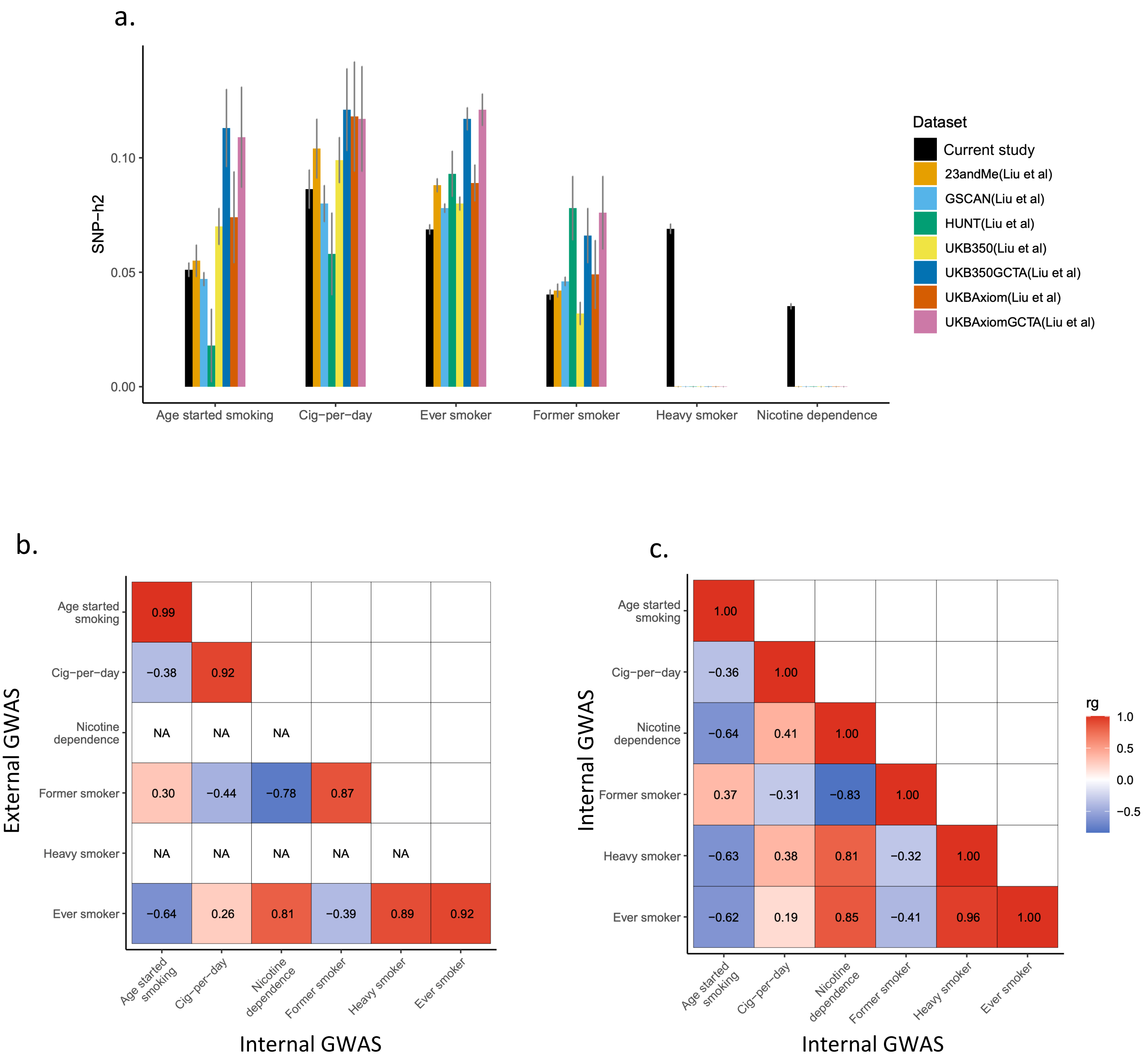


**Supplementary Figure 13. SNP-h2 and genetic correlations.** a. SNP-h2 of the six smoking phenotypes estimated using LD score regression are plotted along with standard errors. The SNP-h2 estimates reported by Liu et al 2019 are also plotted along side for comparisons. Note, Liu et al did not study heavy smoker and nicotine dependence phenotypes. b. Genetic correlations between our six smoking phenotypes and the four smoking phenotypes from GSCAN consortium (Liu et al 2019) estimated using LD score regression are shown as heat maps. The rg estimates are displayed over the plots. Nicotine dependence and heavy smoker phenotypes are not studied by Liu et al, hence, shown as NA. c. Genetic correlations between the smoking phenotypes within our cohort are displayed.

Internal GWAS – Cohorts involved in the current study

External GWAS – GSCAN cohorts (excluding UKB)

a. Ever smoker

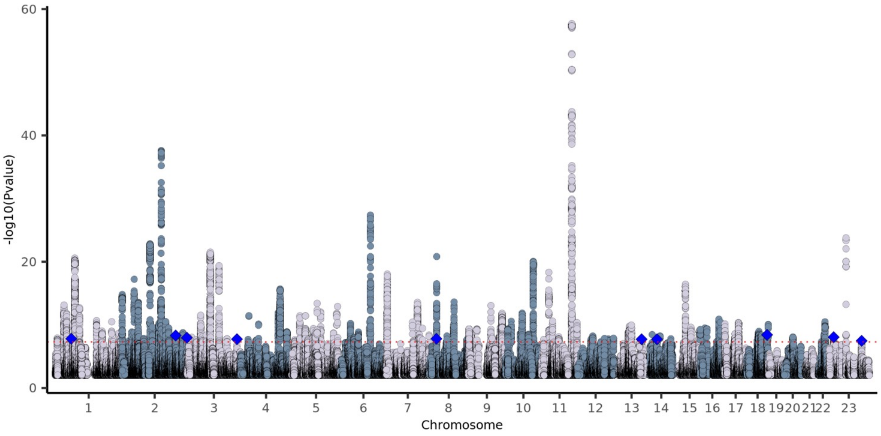


b. Heavy smoker

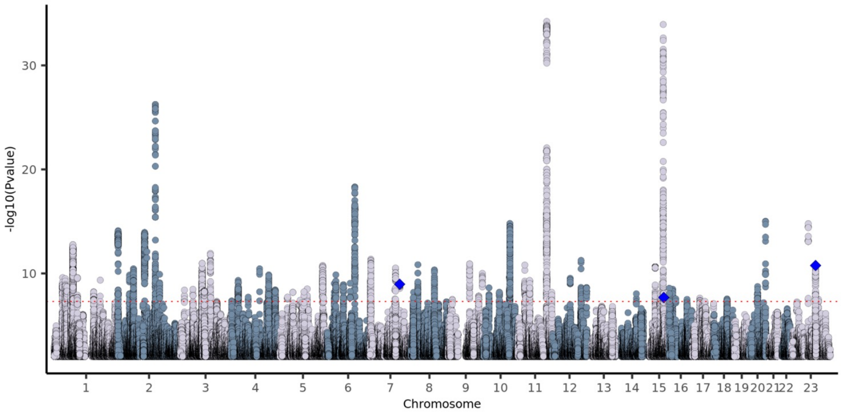


c. Former smoker

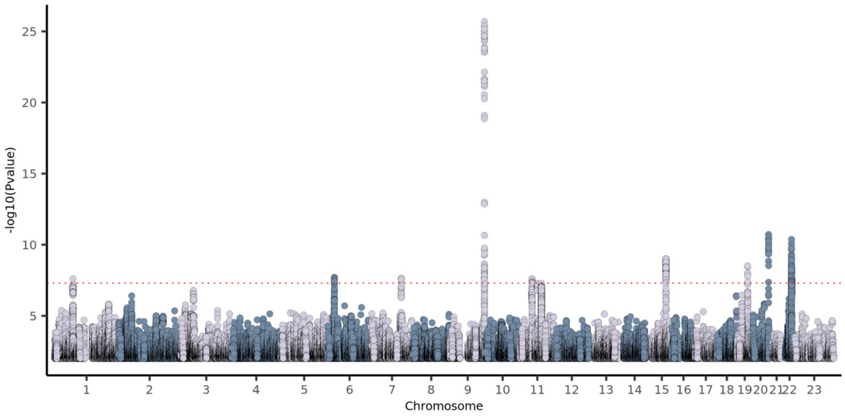

d. Nicotine dependence

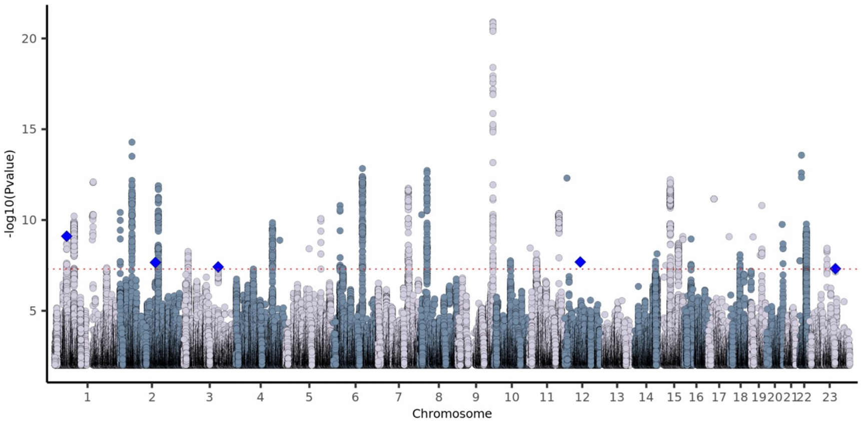


e. Cig-per-day

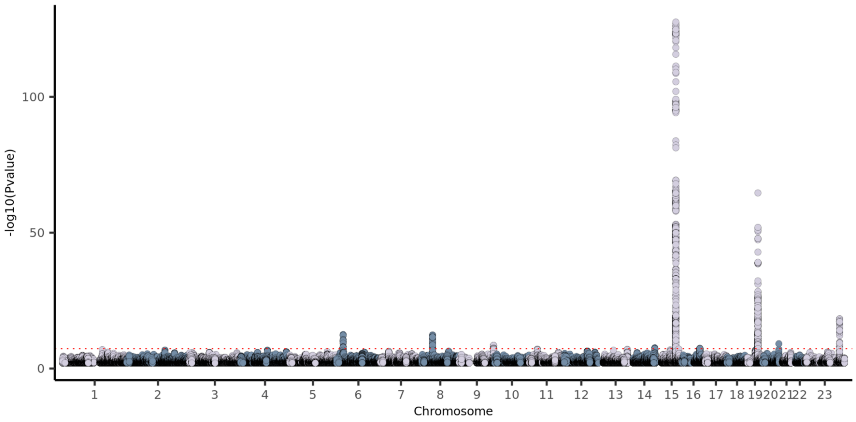


f. Age started smoking

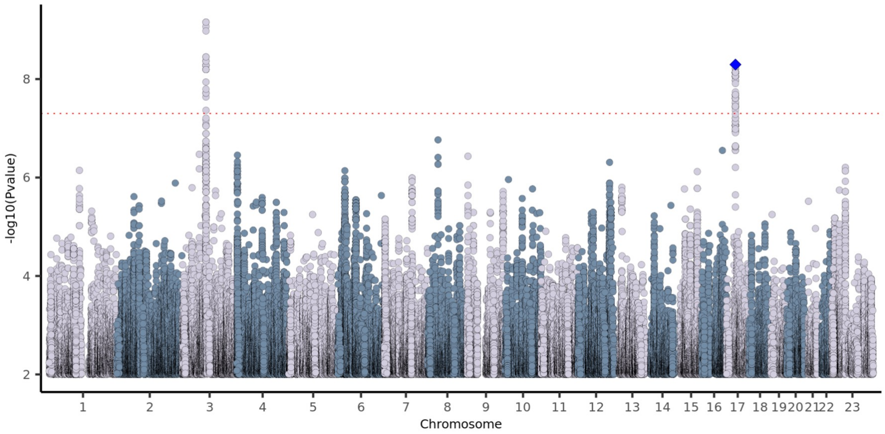


**Supplementary Figure. 14** Manhattan plots of cross-ancestry GWAS meta-analyses of the six smoking phenotypes (a-f). The novel GWAS loci are marked with blue diamonds in the Manhattan plots. The red dotted line corresponds to P value of 5e-8.

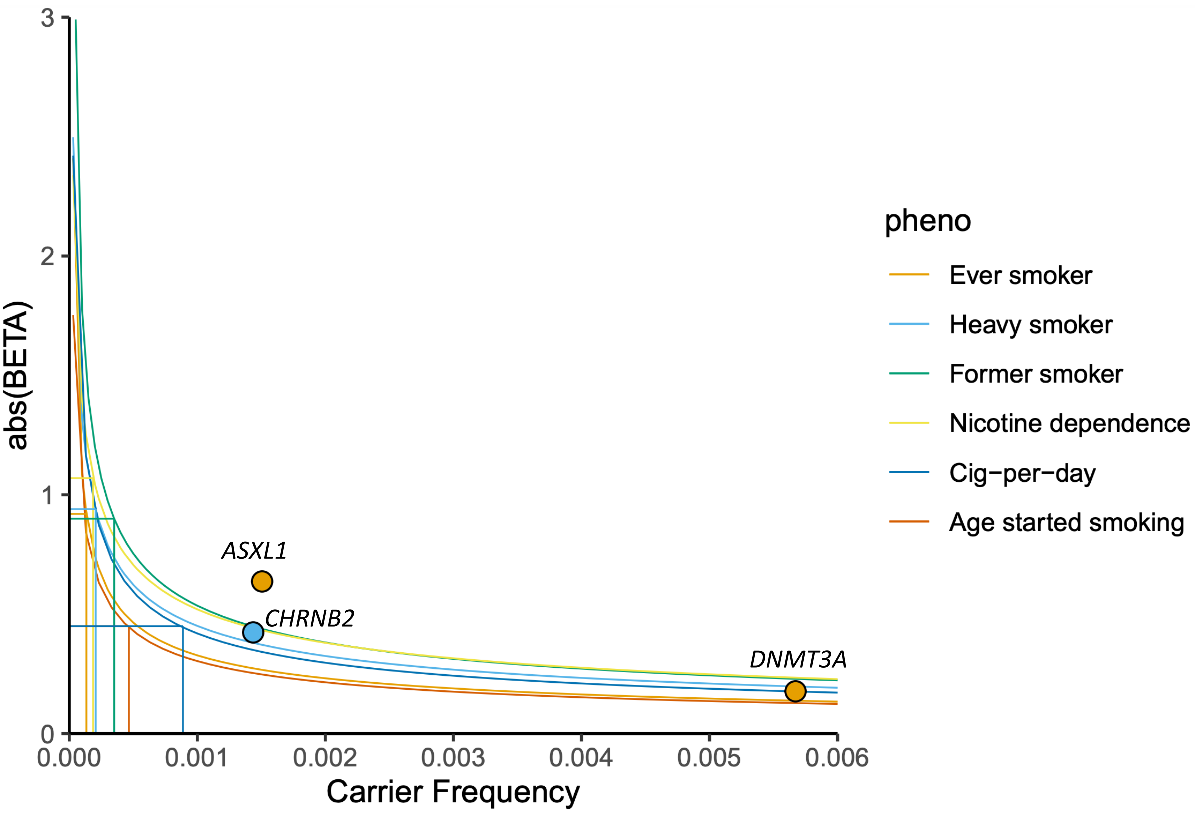


**Supplementary Figure 15. Power calculations for gene discovery using the current sample size.** Assuming an 80% power, P value threshold of 4e-8 (exome-wide significant threshold of the current study based on FDR 1%), effect sizes (i.e., beta values) were computed for a range of minor allele frequencies (combined allele frequency in case of burden masks) for a given sample size (varies across phenotypes). The computed effect sizes (absolute values of beta estimates) are plotted against minor allele frequencies (carrier frequency) for six smoking phenotypes. The carrier frequency corresponding to 100 carriers, calculated for each of the phenotype based on the corresponding sample size, in the X axis and the corresponding effect size in the Y axis are marked with straight lines. The top association of the three genes identified as exome-wide significant are plotted with the color corresponding to the associated phenotype. Based on these power curves, we had 80% power to detect any variant or burden associations with ever smoker, heavy smoker and former smoker with odds ratio ~2.5 or higher (0.4 or lower) when there are at least 100 carriers. And we had 80% power to detect any variant or burden associations with cig-per-day and age started smoking with beta 0.45 (equivalent to 4.7 extra cigarettes for cig-per-day and 1.9 earlier age for age started smoking) when there are at least 100 carriers. These calculations assume that there is no heterogeneity in the effect sizes across the cohorts, which is never the case for complex traits such as smoking. Hence, these estimates should be considered arbitrary. Importantly, the effect sizes for protective associations with binary phenotypes are likely overestimated due to imbalance in the case-control ratios.
